# Predicting cognitive-behavioral therapy outcomes in obsessive-compulsive disorder from inhibitory control neural activity: A mega-analysis and machine learning study from the ENIGMA-OCD consortium

**DOI:** 10.64898/2026.03.13.26348316

**Authors:** Nadža Džinalija, Odile A. van den Heuvel, H. Blair Simpson, Iliyan Ivanov, Pino Alonso, Sara Bertolin, Willem Bruin, Lydia Fortea, Miquel A. Fullana, Kristen Hagen, Bjarne Hansen, Chaim Huijser, Gerd Kvale, Ignacio Martinez-Zalacain, Jose M. Menchon, Olga Therese Ousdal, Carles Soriano-Mas, Anouk L. van der Straten, Sophia I. Thomopoulos, Anders L. Thorsen, Enric Vilajosana, ENIGMA-OCD Consortium, Dan J. Stein, Paul M. Thompson, Ilya M. Veer, Chris Vriend, Laurens A. van de Mortel

## Abstract

**Objective:** Cognitive behavioral therapy (CBT) is an effective first-line treatment for obsessive-compulsive disorder (OCD), yet it remains difficult to predict who will respond to this intervention. This study investigates associations between neural activity during inhibitory control tasks and CBT outcomes, and whether task-based fMRI data could serve as a predictive marker of individual CBT response.

**Methods:** Using fMRI data from individuals performing an inhibitory control task across five samples (n=130, age range=8-57, 54% female) of the ENIGMA-OCD consortium, univariate associations were analyzed between activity during response inhibition and error processing and three CBT outcomes: response, remission, and pre-post treatment change in symptom severity. Random forest and support vector machine models using leave-one-sample-out cross-validation were used for prediction of CBT response and remission from fMRI activity and clinical data.

**Results:** Remission after CBT was associated with weaker activity in default mode regions during response inhibition and in the right supramarginal gyrus during error processing.

Greater symptom reduction was linked to weaker pre-treatment activity across frontoparietal, dorsal attention, visual, and subcortical regions during response inhibition, but to stronger default mode activity during error processing. Despite these robust group-level effects, machine learning models failed to predict individual outcomes above chance level with either neuroimaging or clinical data.

**Conclusion:** Weaker activity during response inhibition in a widespread network, as well as stronger activity in default mode regions during error processing before treatment, appear beneficial to CBT response. However, these findings cannot yet be translated into individually predictive markers of CBT outcome.

## Introduction

Obsessive-compulsive disorder (OCD) affects 2-3% of the general population and is characterized by a pattern of intrusive, distressing thoughts (obsessions) and subsequent anxiety-reducing repetitive behaviors (compulsions) that often runs a chronic and debilitating course (1). Cognitive behavioral therapy (CBT) is a first-line treatment available to individuals suffering from OCD (2), with the most empirically-supported form being exposure with response prevention (ERP; (3)). ERP involves confronting obsession-related stimuli (like a contaminated surface) while refraining from performing compulsions that would normally reduce distress (4). Although CBT is effective for roughly 60% of individuals (5), it is associated with long waiting lists (6) and a considerable burden to the individual (7). These limitations highlight the need to identify predictors of treatment outcome to best target and possibly prioritize those more likely to benefit.

Early efforts to predict treatment response with clinical data have suggested links between poorer outcomes and higher pre-treatment symptom severity, sexual/religious OCD subtypes, and depressive or anxious comorbidities (8–10). However, these findings have been inconsistent.

Resting-state functional connectivity assessed by functional magnetic resonance imaging (fMRI) reflects intrinsic patterns of brain organization that may provide more objective predictors of CBT outcome than clinical data. Greater pre-treatment connectivity in temporal regions within the language network has been linked to larger symptom reduction in OCD patients (11), and increased connectivity between the dorsolateral prefrontal cortex and the precuneus, a key node of the default mode network (DMN), has similarly been associated with greater symptom reduction after CBT (12). However, these statistical associations between brain functional connectivity and treatment outcome are found at the group-level, lacking out-of-sample validation and limiting translation to the individual level.

Machine learning analyses with cross-validation methods offer a way to move from group-level differences towards generalizable and personalized outcome prediction. One study utilizing machine learning showed that resting-state functional connectivity within the default mode and visual networks can predict post-CBT symptom severity more accurately than pretreatment symptom severity (13). However, this finding was not replicated in an ENIGMA-OCD study where resting-state connectivity failed to predict treatment response above chance level, and was outperformed by models based only on clinical data (14).

It may be that task-based fMRI during OCD relevant cognitive processes is more sensitive for predicting CBT treatment response in OCD. Cognitive control processes are a particularly promising candidate, with inhibitory control showing associations with CBT efficacy across psychiatric disorders including depression (15) and PTSD (16). In OCD, greater pre-treatment activity of the anterior cingulate cortex and temporal lobe during goal-directed inhibitory behavior was associated with better CBT response (17). Response inhibition is relevant in OCD because compulsions reflect failures to inhibit repetitive behaviors, and CBT may act partly by enhancing inhibitory control mechanisms. In tasks that tap into these mechanisms, lower anterior insula activity and connectivity during inhibition have been linked to symptom reduction following combined CBT and repetitive transcranial magnetic stimulation (18), whereas higher activity of the sensorimotor network during error processing has been associated with symptom reduction after CBT alone (19). However, these group-level associations may not directly translate to predictive power at the individual patient level. To assess this, machine learning techniques with independent cross-validation are required, but no study has yet applied machine learning to predict CBT outcomes from task-based brain activity in OCD.

In this study, we investigated the association and predictive power of pre-treatment brain activity during inhibitory control tasks on the outcome of CBT for OCD. Using individual participant task-based fMRI data from 130 individuals across five samples of the Enhancing Neuro-Imaging and Genetics through Meta-Analysis (ENIGMA) OCD consortium, we examined differences in neural activity during response inhibition and error processing between different CBT outcome groups, and applied machine learning to clinical and task-based fMRI data to predict individual treatment success. We hypothesized that activation in regions involved in inhibitory control and error processing would be linked to CBT outcomes and support individualized prediction of treatment outcome.

## Methods

### Study population

Five samples were obtained from the ENIGMA-OCD consortium, a global network of institutes with legacy neuroimaging data from OCD individuals. A total of 130 participants (48 pediatric, 82 adults) were identified that had completed a course of CBT (with post-treatment clinical assessment) and had undergone a pre-treatment fMRI scan while performing an inhibitory control task directly prior to the start of CBT treatment (Table 1). OCD diagnosis was determined by DSM-IV or DSM-5 criteria using diagnostic tools administered by trained personnel, while severity was measured using the (Children’s) Yale-Brown Obsessive-Compulsive Scale ((C)Y-BOCS; (20, 21)). Participants gave informed consent at each participating site and protocols were approved by local review boards, who permitted the use of de-identified participant data by ENIGMA-OCD. The present sample was drawn from a larger dataset previously analyzed in Dzinalija et al. (22).

**Table 1.** Demographics of included participants. Values denote mean and standard deviation (SD) unless otherwise stated. Medication status was measured at the time of scan and included any psychoactive medication. (C)Y-BOCS = (Children’s) Yale-Brown Obsessive-Compulsive Scale.

|  | **OCD (n=130)** | **Statistic** |
| --- | --- | --- |
| **Sex** |  |  |
| n female (%) | 70 (53.85%) | - |
| n male | 60 (46.15%) | - |
| **Age** | 24.49 (11.79) | - |
| **Age of onset** |  | - |
| n onset <18 (%) | 92 (70.77%) | - |
| n onset >18 | 35 (26.92%) | - |
| missing data | 3 (2.31%) | - |
| **Medication** |  | - |
| n medicated | 32 (24.62%) | - |
| n unmedicated | 97 (74.62%) | - |
| missing data | 1 (0.77%) | - |
| **(C)Y-BOCS pre CBT** | 25.45 (4.89) | $T_{(129)} = 17.28, p < 0.001$ |
| **(C)Y-BOCS post CBT** | 14.03 (7.65) |  |
| **n responders (%)** | 88 (67.69%) | - |
| **Remitters** | 57 (43.85%) | - |

### CBT treatment

CBT was administered at each site according to slightly different protocols, in individual or group sessions (Table S2). Participants completed between 16 and 24 hours of CBT over a period of 1-27 weeks (average 9.7 weeks). Treatment outcome was operationalized in three ways. First, treatment response was defined categorically as a reduction of ≥35% in (C)Y-BOCS score from pre- to post- treatment (22). Second, remission was defined as attaining a post-treatment (C)Y-BOCS score <=12, indicating OCD symptoms diminished to subclinical levels (23). Finally, treatment outcome was quantified continuously as the absolute change in (C)Y-BOCS score between pre-treatment minus post-treatment score.

### Inhibitory control tasks

Participants from each sample completed an inhibitory control task in the MRI scanner (Table 2) consisting of either a stop-signal (four samples) or a flanker task (one sample). Stop-signal tasks assessed action cancellation, i.e., the inhibition of an already initiated motor response, while the flanker task assessed interference control, i.e., the suppression of intrusions by distracting or irrelevant stimuli. Prior work (22) using the same flanker and stop-signal tasks demonstrated highly similar activation patterns in the main task effect across OCD and healthy control participants, thereby supporting our decision to pool these tasks in the current analyses.

**Table 2.** fMRI inhibitory control tasks. All tasks modeled events using event-related regressors with zero duration (i.e., stick functions)

| Site PI | City,<br>Country | Task | Inhibition<br>events | Response<br>events | INHIBITION<br>contrast | ERROR<br>contrast | n OCD<br>(age<br>range) |
| --- | --- | --- | --- | --- | --- | --- | --- |
| Huyser (53) | Amsterdam,<br>NLD | Flanker | Incongruent | Congruent | Incong. > Cong. | Incong. fail ><br>Incong. success | 27<br>(8-19) |
| van Wingen (54) | Amsterdam,<br>NLD | Stop-signal | Stop | Go | Stop success ><br>Go | Stop fail ><br>Stop success | 14<br>(20-55) |
| Fullana (55) | Barcelona,<br>ESP | Stop-signal | Stop | Go | Stop success ><br>Go | Stop fail ><br>Stop success | 31<br>(9-47) |
| Menchón/<br>Soriano-Mas (56) | Barcelona,<br>ESP | Stop-signal | Stop | Go | Stop success ><br>Go | Stop fail ><br>Stop success | 22<br>(22-57) |
| Thorsen (57) | Bergen,<br>NOR | Stop-signal | Stop | Go | Stop success ><br>Go | Stop fail ><br>Stop success | 36<br>(19-54) |

### MRI Image acquisition and processing

MRI data acquisition and preprocessing followed identical procedures to those described in Dzinalija et al. (22). All imaging data were previously acquired across multiple sites (see Table S3 for acquisition parameters). Image processing was harmonized using the open-source containerized Harmonized AnaLysis of Functional MRI pipeline (HALFpipe; version 1.2.2; (24)), using default settings (see Supplement). For structural images this included skull stripping, tissue segmentation, and spatial normalization, and for functional images included motion correction and calculation of six rigid-body motion parameters, slice time correction (if slice acquisition order was known), susceptibility distortion correction (if fieldmaps were available), coregistration, spatial normalization, high-pass filtering, denoising with six (rigid-body) motion parameters, and smoothing with a 6mm FWHM Gaussian kernel. Participants with mean framewise displacement >1.0mm were excluded and processed data was quality controlled at each site according to harmonized guidelines (see Supplement).

### First-level contrast parameter estimate maps

We focused on two participant-level task contrasts available across datasets: response inhibition and error processing. The response inhibition contrast compared trials that require the inhibition/suppression of a response (stop trials in the Stop-signal tasks and incongruent trials in the Flanker task) with response conditions (go trials in the Stop-signal tasks and congruent trials in the Flanker task) (Table 1). The error contrast modeled commission errors, when an erroneous response was made to a stop or incongruent trial, and contrasted them with successful inhibition trials. For participants with multiple runs, individual first-level contrast parameter estimate maps were averaged across runs to obtain a single contrast map per participant.

### Analyses

#### ROI and whole-brain activity measures

We investigated regions of interest (ROIs) previously implicated in inhibitory control and error processing, based on coordinates reported in the meta-analysis by Norman et al. (25) (Table S4). Spherical ROIs (5 mm radius) were constructed around each coordinate in MNI152 NLIN 2009c (asymmetric) space, resulting in 12 ROIs for response inhibition and 7 for error processing. For each participant, we extracted the mean activity within each ROI from the first-level z-statistic maps corresponding to the response inhibition and error processing contrasts.

In addition to these a priori ROIs, we conducted a whole-brain analysis using functionally defined anatomical parcels. Cortical activity was extracted using the 200-parcel, Schaefer-Yeo 7-network atlas (26), and subcortical activity was extracted using scale 2 of the Melbourne subcortical atlas (27). As for the ROIs, mean activity within each parcel was computed from the first-level z-statistic maps, provided 30% of voxels contained signal (see Table S5 for exclusions).

#### Univariate analyses

We assessed whether pre-treatment task-based brain activity was associated with the three clinical outcomes using mixed-effects models. The outcome for CBT (response, remission, and absolute change in symptom severity) were examined separately for the response inhibition and error processing contrasts. All models included age and sex as covariates of no interest. For analyses examining associations with symptom change, pre-treatment (C)Y-BOCS score was also included as a covariate to account for pre-treatment differences between patients. Sample/scanner effects were removed using ComBat as implemented in the sva R package (v3.46.0) (28), preserving variation associated with the outcome variable as well as age, sex, and when relevant, pre-treatment (C)Y-BOCS score. Statistical models were fit at the group level using linear mixed-effects models. Each ROI or parcel was analyzed separately, modeling pre-treatment activity as a function of the clinical outcome variables. Multiple comparison correction was applied with false discovery rate (FDR) correction at α = 0.05. To assess pre-treatment differences between groups, clinical variables were compared between responders and non-responders using two-sample t-tests, chi-square tests, or Wilcoxon rank-sum tests where appropriate at α = 0.05 and FDR-corrected for multiple comparisons. All univariate analyses were conducted in R (v. 4.3.2; (29)).

#### Sensitivity Analyses

To assess the robustness of our findings, we conducted four sensitivity analyses. First, to compare different sample correction methods, we compared results from ComBat-corrected models with mixed-effects models including a random intercept for sample. Second, to account for within-participant correlations across ROIs, we fit a combined mixed-effects model including all ROIs simultaneously, with random intercepts for both participant and sample. Third, we repeated the main analyses including medication status as an additional covariate. Finally, we conducted parallel Bayesian multilevel analyses using the RBA package (v1.0.10; (30)). Details of the Bayesian models are provided in the Supplement.

#### Machine learning analyses

Binary machine learning classification of CBT response and remission was performed by a leave-one-site-out cross-validation loop. Performance for both random forest (RF) and support vector machine (SVM) models on CBT outcome prediction were compared for the use of mean ROI activity, whole-brain activity, and clinical features. Clinical features consisted of the participants’ age, biological sex, age of onset (categorical, <18 or ≥18 years), current medication status (categorical, yes/no), and pre-treatment (C)Y-BOCS.

For each training instance, an inner 5-fold grid search was performed on the training data to find optimal hyperparameters for SVM (C: 0.1-1000, gamma: 0.0001-1, linear or radial basis function kernel) and RF (max features: 10-300, minimum samples per leaf: 1-10, minimum samples per split: 2-20, number of estimators: 100-1000) with balanced accuracy as the scoring function. The inner folds were stratified on CBT outcome and Z-scaling of the features was done on the training and test data separately prior to the grid search.

Besides the leave-one-site-out cross validation, we performed an additional analysis using outcome-stratified nested 5x5 cross-validation with shuffled data across all samples. To mitigate noise and bias affecting the learning of relevant features in multi-site data, a ComBat regressor was trained on the training data to estimate and harmonize effects of sample/scanner and subsequently applied to the testing data.

Over all folds, we averaged the area under the receiver-operating-characteristic curve (AUC), sensitivity, specificity, positive predictive value (PPV), and negative predictive value (NPV) and calculated corresponding 95% confidence intervals for the AUC values using an analytical computation of the DeLong method (31). Statistical significance of machine learning model performance were assessed with 100 permutations when lower bounds of the AUC confidence intervals exceeded 0.5.

## Results

### Participants

The sample included 130 participants (53.9% female, mean age=24.5±11.8, 74.6% unmedicated) (Table 1,S1), with a pre-treatment mean (C)Y-BOCS score of 25.5±4.9, indicating moderate-to-severe OCD symptoms. Following CBT, mean (C)Y-BOCS score decreased to 14.0±7.7, representing a significant decrease in severity (T_(129)_=17.28, p<0.001). Overall, 67.7% of participants met the criterion for treatment response, while 43.9% achieved remission.

### Univariate analyses

### Response

Response to CBT was not associated with pre-treatment task-based activity in any whole-brain parcel or ROI, neither during response inhibition nor error processing. There were no significant differences in baseline clinical variables between responders and non-responders.

### Remission

CBT remission was associated with weaker inhibition-related activity in the precuneus/posterior cingulate (pCunPCC) regions of the DMN (left pCunPCC 3: T_(126)_=-3.784, p_FDR corr._=0.028; right pCunPCC 2: T_(126)_=-3.826, p_FDR corr._=0.028) and weaker error-related activity in the right supramarginal gyrus (SMG; T_(126)_=-2.756, p_FDR corr._=0.047). Remitters showed lower pre-treatment (C)Y-BOCS score (M=23.9±5.4) than non-remitters (M=26.7±4.0), T_(101)_=3.2, p=0.002).

### Symptom change

Regression analyses revealed a negative association between change in (C)Y-BOCS following CBT and pre-treatment inhibition-related activity in the right primary motor cortex (PMC), bilateral supplementary motor area (SMA), and right superior parietal lobule (SPL) in ROI analyses. In the whole-brain analyses, significant negative associations also emerged across multiple regions spanning DMN (medial and lateral prefrontal cortex, precuneus / posterior cingulate, inferior parietal lobule), frontoparietal network (dorsolateral and ventrolateral prefrontal cortex, parietal cortex, anterior cingulate), dorsal attention network (posterior parietal, occipital cortex), visual cortex, thalamus, and posterior hippocampus (all p_FDR corr._<0.05; Table S6). There was also a positive association between change in (C)Y-BOCS and error-processing activity in precuneus/posterior cingulate regions of the DMN (left pCunPCC 2: T_(125)_=3.855, p_FDR corr._=0.04; right pCunPCC 2: T_(125)_=3.677, p_FDR corr._=0.04).

### Sensitivity analyses

Sensitivity analyses with alternative mixed-effects models - using a random intercept for sample, modelling all ROIs simultaneously with random intercepts for participant and sample, or adding medication status as a covariate – yielded results broadly consistent with the main ComBat-harmonized analyses (Figures S1,S2; Tables S7). Across analyses, modelling all ROIs simultaneously was more sensitive to differences between CBT outcome groups. Bayesian multilevel models provided greater sensitivity, detecting effects that the frequentist models did not (Figures S3-7). For response inhibition, CBT responders showed credible evidence (i.e., the posterior probability strongly supported an effect) for weaker pre-treatment inhibition-related activity than non-responders across several ROI and whole-brain regions. For error processing, CBT remission was associated with weaker pre-treatment error-related activity in all ROIs. Whole-brain Bayesian analyses of error processing revealed more widespread effects than frequentist analyses, with credible evidence for stronger error-related activity in temporal, default mode, and prefrontal regions in both responders and those with larger symptom reduction.

### Machine learning

The best predictive performance of CBT outcome was achieved by a SVM using 5-fold cross-validation for the prediction of treatment response from ROI activity during response inhibition (AUC=0.59), but this was not statistically significant (p>0.05). Other average performance metrics of the machine learning models did not exceed chance-level for both leave-one-site-out and 5-fold cross-validation methods (Figure 2). These chance-level results were obtained for both neuroimaging and clinical data and for the prediction of both CBT response and remission, with large variation in prediction performance and confidence intervals over the different folds (Table 3).

**Figure 1.**
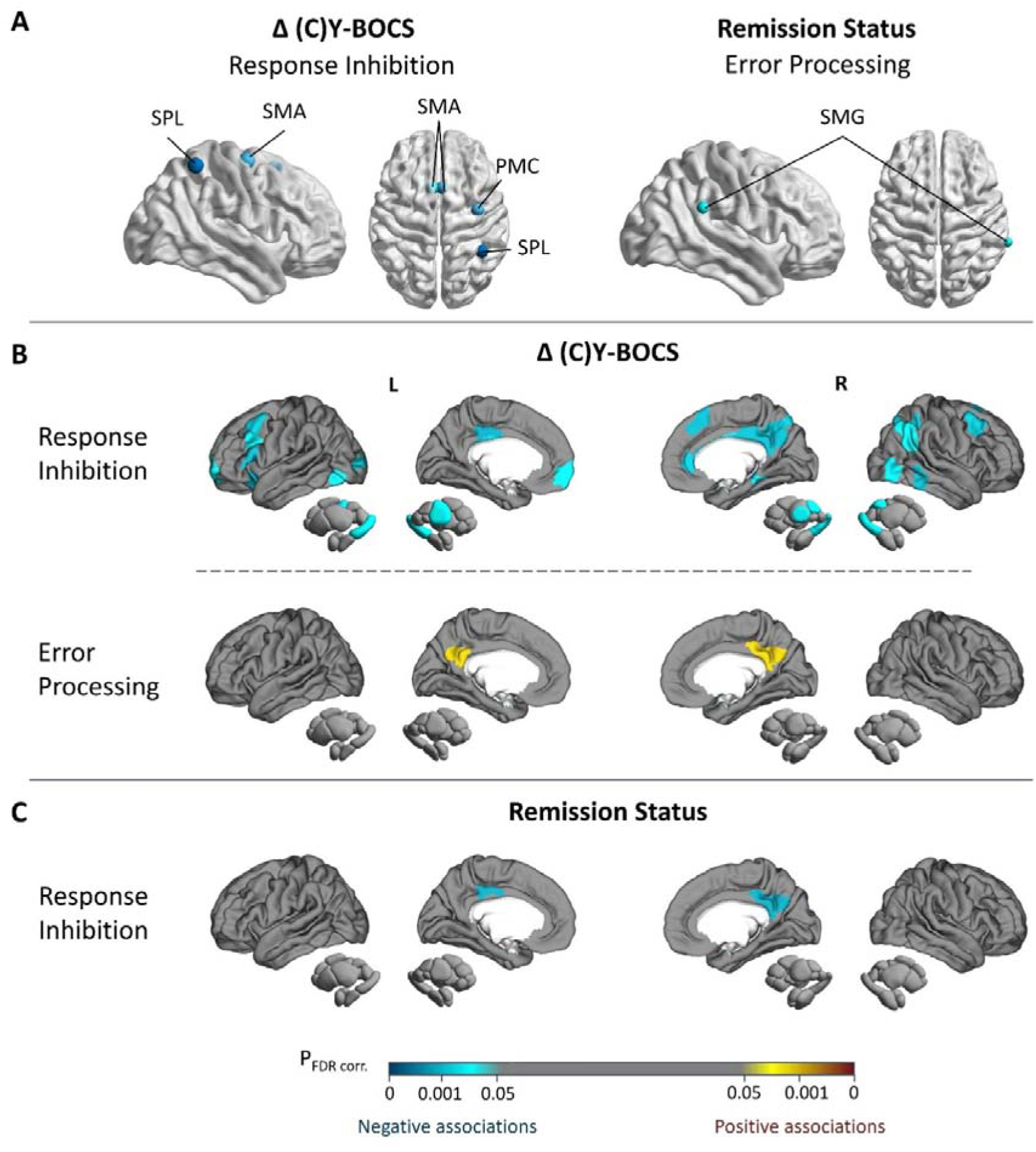
Brain activity during response inhibition and error processing and association with CBT outcomes. Results of univariate analyses relating brain activity during response inhibition and error processing to change in (C)Y-BOCS score and remission. (A) Region of interest analysis (B,C) Whole brain analysis. Regions are color coded to reflect significance of effect after FDR correction. Full reporting of all investigated effects can be found in Table S6. Displayed are lateral and medial views of the cortex and subcortex (amygdala, hippocampus, thalamus, nucleus accumbens, pallidum, putamen, and caudate). Δ (C)Y-BOCS = change in (Children’s) Yale-Brown Obsessive-Compulsive Scale total score (pre-treatment minus post-treatment); L = left; FDR = false discovery rate; PMC = primary motor cortex; R = right; SMA = supplementary motor area; SMG = supramarginal gyrus; SPL = superior parietal lobule.

**Figure 2.**
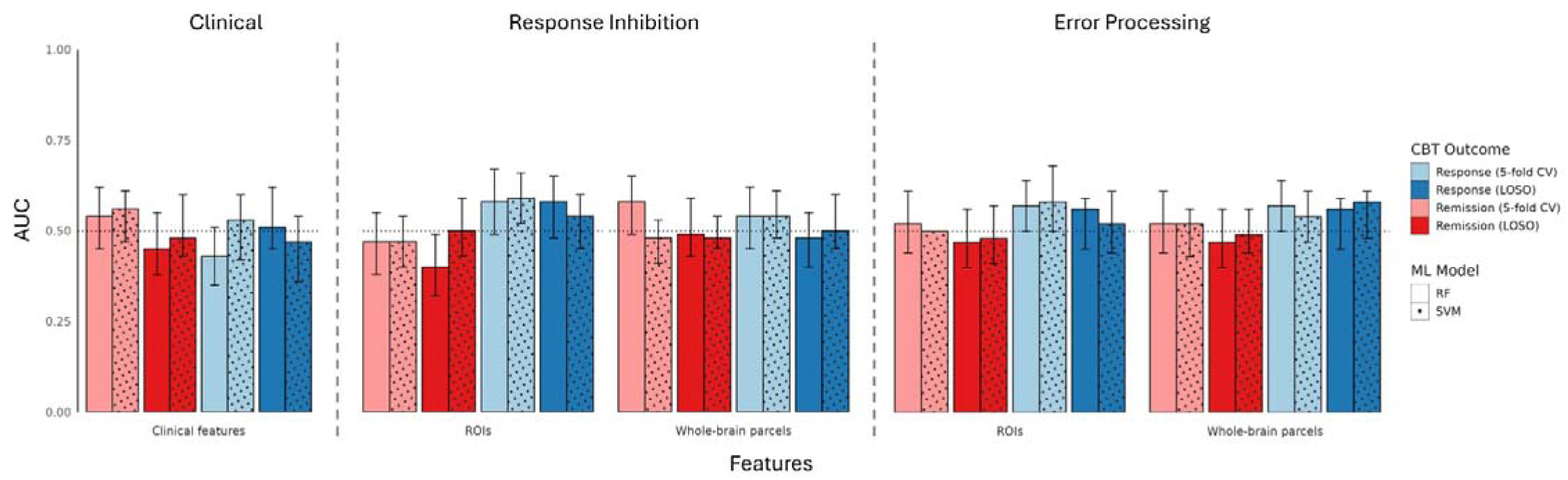
Machine learning model performance of predicting CBT outcome. Men AUC values for prediction models trained on clinical data (age, biological sex, age of onset, medication status, symptom severity) as well as response inhibition and error processing ROI and whole-brain activity data. Models for each CBT outcome, cross-validation method, and machine learning model are shown. Error bars denote confidence intervals. AUC = area under the receiver-operating-characteristic curve; CBT = Cognitive Behavioral Therapy; CV = cross-validation; LOSO = leave-one-site-out; ML = machine learning; RF = random forest, ROIs = regions of interest; SVM = support vector machine.

**Table 3.** Results of machine learning analyses. Classification performance when predicting CBT response and remission using neural activity data during response inhibition and error processing, as well as using only clinical data (age, biological sex, age of onset, medication status, symptom severity). Regions of interest (ROIs) refer to predefined spheres around coordinates reported in Norman et al. (7) for response inhibition and error processing. Whole-brain parcels refer to the Schaefer 200-parcel 7-network cortical atlas (5) and Melbourne 32-region subcortical atlas (6). ROIs refer to AUC = area under the receiver operating curve; CI = confidence interval; CV = cross validation; LOSO = leave-one-site-out; NPV = negative prediction value; PPV = positive prediction value.

| Model | Features | Outcome | Cross-validation | AUC (CI) | Sensitivity (SD) | Specificity (SD) | PPV (SD) | NPV (SD) |
| --- | --- | --- | --- | --- | --- | --- | --- | --- |
| **Response Inhibition** |  |  |  |  |  |  |  |  |
| SVM | ROIs | Response | LOSO | 0.54 (0.45-0.60) | 0.27 (0.24) | 0.80 (0.08) | 0.66 (0.10) | 0.38 (0.19) |
| SVM | ROIs | Remission | LOSO | 0.50 (0.43-0.59) | 0.30 (0.11) | 0.70 (0.13) | 0.43 (0.13) | 0.57 (0.12) |
| SVM | ROIs | Response | 5-fold CV+ComBat | 0.59 (0.52-0.66) | 0.32 (0.13) | 0.86 (0.08) | 0.84 (0.09) | 0.38 (0.09) |
| SVM | ROIs | Remission | 5-fold CV+ComBat | 0.47 (0.40-0.54) | 0.15 (0.09) | 0.79 (0.12) | 0.34 (0.25) | 0.54 (0.08) |
| Random forest | ROIs | Response | LOSO | 0.58 (0.48-0.65) | 0.76 (0.13) | 0.40 (0.10) | 0.71 (0.14) | 0.50 (0.24) |
| Random forest | ROIs | Remission | LOSO | 0.40 (0.32-0.49) | 0.30 (0.11) | 0.50 (0.11) | 0.31 (0.11) | 0.49 (0.11) |
| Random forest | ROIs | Response | 5-fold CV+ComBat | 0.58 (0.49-0.67) | 0.70 (0.12) | 0.46 (0.14) | 0.73 (0.10) | 0.43 (0.04) |
| Random forest | ROIs | Remission | 5-fold CV+ComBat | 0.47 (0.38-0.55) | 0.39 (0.10) | 0.54 (0.23) | 0.44 (0.22) | 0.51 (0.14) |
| SVM | Whole-brain parcels | Response | LOSO | 0.50 (0.45-0.60) | 0.20 (0.19) | 0.81 (0.13) | 0.48 (0.41) | 0.34 (0.13) |
| SVM | Whole-brain parcels | Remission | LOSO | 0.48 (0.45-0.54) | 0.07 (0.13) | 0.90 (0.09) | 0.13 (0.27) | 0.57 (0.10) |
| SVM | Whole-brain parcels | Response | 5-fold CV+ComBat | 0.54 (0.48-0.61) | 0.22 (0.05) | 0.87 (0.14) | 0.84 (0.16) | 0.35 (0.08) |
| SVM | Whole-brain parcels | Remission | 5-fold CV+ComBat | 0.48 (0.41-0.53) | 0.13 (0.12) | 0.84 (0.11) | 0.38 (0.38) | 0.55 (0.12) |
| Random forest | Whole-brain parcels | Response | LOSO | 0.48 (0.40-0.55) | 0.70 (0.26) | 0.26 (0.21) | 0.61 (0.18) | 0.24 (0.14) |
| Random forest | Whole-brain parcels | Remission | LOSO | 0.49 (0.43-0.59) | 0.22 (0.22) | 0.75 (0.15) | 0.30 (0.30) | 0.57 (0.07) |
| Random forest | Whole-brain parcels | Response | 5-fold CV+ComBat | 0.54 (0.45-0.62) | 0.74 (0.09) | 0.34 (0.16) | 0.70 (0.09) | 0.37 (0.10) |
| Random forest | Whole-brain parcels | Remission | 5-fold CV+ComBat | 0.58 (0.49-0.65) | 0.47 (0.06) | 0.70 (0.20) | 0.58 (0.24) | 0.61 (0.11) |
| **Error Processing** |  |  |  |  |  |  |  |  |
| SVM | ROIs | Response | LOSO | 0.52 (0.44-0.61) | 0.34 (0.20) | 0.71 (0.14) | 0.68 (0.09) | 0.36 (0.19) |
| SVM | ROIs | Remission | LOSO | 0.48 (0.41-0.57) | 0.24 (0.16) | 0.72 (0.13) | 0.38 (0.17) | 0.56 (0.12) |
| SVM | ROIs | Response | 5-fold CV+ComBat | 0.58 (0.50-0.68) | 0.52 (0.23) | 0.64 (0.06) | 0.70 (0.17) | 0.41 (0.08) |
| SVM | ROIs | Remission | 5-fold CV+ComBat | 0.50 (0.44-0.56) | 0.13 (0.13) | 0.86 (0.15) | 0.37 (0.37) | 0.56 (0.10) |
| Random forest | ROIs | Response | LOSO | 0.56 (0.45-0.59) | 0.91 (0.11) | 0.21 (0.18) | 0.69 (0.16) | 0.47 (0.38) |
| Random forest | ROIs | Remission | LOSO | 0.47 (0.40-0.56) | 0.21 (0.18) | 0.72 (0.15) | 0.36 (0.27) | 0.56 (0.07) |
| Random forest | ROIs | Response | 5-fold CV+ComBat | 0.57 (0.50-0.64) | 0.92 (0.06) | 0.22 (0.08) | 0.71 (0.07) | 0.63 (0.19) |
| Random forest | ROIs | Remission | 5-fold CV+ComBat | 0.52 (0.44-0.61) | 0.35 (0.13) | 0.69 (0.26) | 0.57 (0.25) | 0.56 (0.15) |
| SVM | Whole-brain parcels | Response | LOSO | 0.58 (0.48-0.61) | 0.23 (0.18) | 0.92 (0.15) | 0.69 (0.41) | 0.38 (0.15) |
| SVM | Whole-brain parcels | Remission | LOSO | 0.49 (0.44-0.56) | 0.10 (0.10) | 0.88 (0.10) | 0.29 (0.30) | 0.57 (0.08) |
| SVM | Whole-brain parcels | Response | 5-fold CV+ComBat | 0.54 (0.47-0.61) | 0.23 (0.09) | 0.85 (0.12) | 0.80 (0.13) | 0.35 (0.10) |
| SVM | Whole-brain parcels | Remission | 5-fold CV+ComBat | 0.52 (0.43-0.56) | 0.18 (0.17) | 0.85 (0.16) | 0.45 (0.33) | 0.57 (0.10) |
| Random forest | Whole-brain parcels | Response | LOSO | 0.56 (0.45-0.59) | 0.91 (0.11) | 0.21 (0.18) | 0.69 (0.16) | 0.47 (0.38) |
| Random forest | Whole-brain parcels | Remission | LOSO | 0.47 (0.40-0.56) | 0.21 (0.18) | 0.72 (0.15) | 0.36 (0.27) | 0.56 (0.07) |
| Random forest | Whole-brain parcels | Response | 5-fold CV+ComBat | 0.57 (0.50-0.64) | 0.92 (0.06) | 0.22 (0.08) | 0.71 (0.07) | 0.63 (0.19) |
| Random forest | Whole-brain parcels | Remission | 5-fold CV+ComBat | 0.52 (0.44-0.61) | 0.35 (0.13) | 0.69 (0.26) | 0.57 (0.25) | 0.56 (0.15) |
| **Clinical Features** |  |  |  |  |  |  |  |  |
| SVM | Clinical features | Response | LOSO | 0.47 (0.36-0.54) | 0.63 (0.31) | 0.30 (0.34) | 0.61 (0.15) | 0.15 (0.15) |
| SVM | Clinical features | Remission | LOSO | 0.48 (0.43-0.60) | 0.32 (0.22) | 0.64 (0.08) | 0.38 (0.24) | 0.57 (0.12) |
| SVM | Clinical features | Response | 5-fold CV+ComBat | 0.53 (0.42-0.60) | 0.56 (0.26) | 0.51 (0.32) | 0.74 (0.13) | 0.28 (0.16) |
| SVM | Clinical features | Remission | 5-fold CV+ComBat | 0.56 (0.47-0.61) | 0.27 (0.19) | 0.84 (0.11) | 0.48 (0.33) | 0.59 (0.12) |
| Random forest | Clinical features | Response | LOSO | 0.51 (0.45-0.62) | 0.79 (0.07) | 0.24 (0.26) | 0.68 (0.15) | 0.32 (0.27) |
| Random forest | Clinical features | Remission | LOSO | 0.45 (0.38-0.55) | 0.50 (0.09) | 0.41 (0.27) | 0.41 (0.14) | 0.43 (0.23) |
| Random forest | Clinical features | Response | 5-fold CV+ComBat | 0.43 (0.35-0.51) | 0.64 (0.07) | 0.22 (0.07) | 0.63 (0.10) | 0.22 (0.03) |
| Random forest | Clinical features | Remission | 5-fold CV+ComBat | 0.54 (0.45-0.62) | 0.48 (0.17) | 0.61 (0.10) | 0.47 (0.15) | 0.60 (0.14) |

## Discussion

In this ENIGMA-OCD study across five different samples, we investigated associations between pre-treatment neural activity during response inhibition and error processing and CBT outcome. We also assessed whether this data could serve as a predictive marker at the individual level. Univariate analyses revealed associations between remission and weaker pre-treatment inhibition-related activity in default mode regions and error-related activity in the right SMG. Larger symptom reduction was associated with weaker pre-treatment inhibition-related activity in a number of ROIs relevant for inhibitory control (including the SPL, PMC and SMA), as well as in widespread frontoparietal, default mode, dorsal attention, hippocampal, thalamic, and visual regions. An opposite pattern was observed during error processing, where stronger pre-treatment default mode activity was associated with greater symptom reduction. Although our reported group differences were detected across sensitivity analyses and were particularly robust in Bayesian analyses, they did not lead to reliable individual prediction of CBT response in machine learning analyses. Over the different cross-validation methods and models, machine learning performance did not exceed chance-level prediction for neither task-based fMRI nor clinical data.

The group-level differences we found suggest that weaker pre-treatment activity during response inhibition is associated with better CBT outcome, a finding which contrasts with several prior studies that favor stronger global pre-treatment activity during inhibitory control tasks (17, 18, 32). While we did not find stronger pre-treatment inhibitory activity associations, we found stronger error-related activity to be associated with symptom reduction. Notably, the regions showing this effect were not canonical error-processing areas, but default mode regions including the precuneus and posterior cingulate cortex which are typically suppressed during task performance (33). This finding aligns with prior work showing stronger error-related activity in overlapping sensorimotor regions in CBT responders (19). Activity in these same sensorimotor regions was found to have a positive correlation with symptom change following CBT in an emotional processing task (34). Enhanced error-related DMN activity may reflect difficulty disengaging from internally oriented processing, a pattern observed in OCD (35) and related anxiety disorders (36, 37). CBT may act in part by improving DMN downregulation in key DMN hubs (including the medial PFC, ACC, and precuneus) (38) and by strengthening frontostriatal cognitive control circuits (39). Supporting the relevance of the DMN in OCD, the cingulate cortex, a region partly overlapping with the DMN, was the most relevant region in discriminating individuals with OCD from healthy controls in machine learning analyses using resting-state functional data (40).

Taken together, these results indicate that greater deviation from typical inhibitory-and error-processing activity relate to higher CBT improvement. In line with our prior work linking reduced inhibitory activity to greater OCD severity (22), this suggests that patients with higher task-based neural activity deviation exhibit greater treatment-related change (19).

The differences in associations we found across treatment outcomes and compared with other studies underscores that the detection of group differences depends strongly on how CBT success is defined: dimensional measures of symptom change revealed the most widespread effects, while a dichotomous definition of treatment response showed none. These associations also depend on which tasks paradigms are used, as different task loads can induce different activity patterns and therefore different directional effects (41). A meta-analysis largely based on emotional processing tasks (such as symptom provocation and fear conditioning) identified stronger pre-treatment activity in the SPL and SMA to be associated with symptom reduction (42), a pattern opposite to that observed here using inhibitory control tasks. A recent machine-learning study using a fear-conditioning task in social anxiety disorder reported promising predictive power (43), suggesting that tasks that probe emotional processes may provide greater sensitivity than inhibitory control alone.

While our group-level findings using task-based fMRI show associations with CBT outcome, these findings did not generalize well to the individual level as indicated by our chance-level machine learning performance in both the task-based fMRI and clinical data. Our inability to translate these group-level differences into meaningful predictors of treatment outcomes is not in line with previous machine learning studies that have succeeded in predicting CBT outcomes using task-based activity in depression (44, 45) and anxiety disorders (43, 46, 47). However, these studies were performed with smaller, homogeneous samples and may therefore limit generalizability to larger, more heterogeneous patient populations or multi-site data (48). In fact, several studies have failed to identify reliable neural predictors of treatment sensitivity using machine learning in larger samples (49, 50), including most recently a resting-state study in ENIGMA-OCD whose sample overlapped partially with ours (14). Associations between brain function and treatment outcome found at the group level, such as those reported in this study, may therefore not reflect markers that are directly relevant to the individual level.

While it is known that group-level associations modelled on the entire sample are not always useful for individual-level predictions on held-out data (51, 52), and that small homogeneous samples in machine learning limit generalizability to a broader clinical population, the discrepancy between group-level and individual results should also be seen in the context of this study’s limitations. First, there was high variation between samples in demographics, task paradigms, scanners, treatment protocols and efficacy. While prior work using the same task paradigms demonstrated highly similar activation patterns supporting their pooling (22), some degree of task-related heterogeneity likely remains. Although a broader range of clinical and demographic variables such as symptom severity, education level, and OCD subtype may generalize relatively well in the prediction of CBT outcome across samples (14), we did not have access to all these variables in our study.

These differences therefore did not lead to individual-level predictability as more influential site-specific factors not captured by our data such as treatment protocols, patient subtypes, or other influences may explain CBT outcome variance better than the clinical or task-based fMRI data. Additionally, while our sample size was relatively large for a clinical neuroimaging study, several hundred participants are recommended to account for high clinical heterogeneity and allow robust predictive modelling (51). Prospective large-scale cohorts should therefore be created in the future to assess whether pretreatment task-based neural activity in OCD may be a sufficiently sensitive predictor of CBT outcome. These cohorts should focus on a better trade-off between sample size and patient variation and a broader range of cognitive and emotional fMRI paradigms.

## Conclusion

This ENIGMA-OCD study highlights the relevance of inhibitory control function in OCD for CBT outcome, with better outcomes related to weaker pre-treatment global inhibition-related activity and stronger error-related activity in the default mode network. However, it is important to recognize the limitations of these findings for the individual patient, as machine learning analyses did not indicate any predictive value of neural activity data during inhibitory control tasks on CBT outcomes. More large-scale studies are required to investigate how these functional associations can be translated to effective personalized treatment of OCD.

## Data Availability

All data produced in the present work are contained in the manuscript.

## Acknowledgements

The ENIGMA-Obsessive Compulsive Disorder Working-Group gratefully acknowledges support from the International Obsessive-Compulsive Disorder Foundation with the Innovator Award of 2021 awarded to OAvdH and CV. The authors were further supported by the Spanish Ministry of Science, Innovation and Universities and Universities Agencia Estatal de Investigación (Grant No. ISCIII PI22/00752 to PA and 10.13039/501100011033 and PID2022-139081OB-C21 to MAL); the Catalan Audiovisual Media Corporation La Fundació Marató de TV3 (Grant No. 202201 to CSM and PA); the Rio Hortega (Grant No. CM21/00278 to SB); the European Regional Development Fund (FEDER); the Carlos III Health Institute (Grant No. PI19/01179 to CSM); the South African Medical Research Council; the National Institute of Health (Grant No. R01MH138569 to PMT, OvdH, HBS); and the Western Norway Health Authority (Grants 911754 and 911880 to ALT).

## References

1. Stein DJ, Costa DLC, Lochner C, Miguel EC, Reddy YCJ, Shavitt RG, et al. Obsessive-compulsive disorder. Nat Rev Dis Primers. 2019;5(1):52.

2. Swierkosz-Lenart K, Dos Santos JFA, Elowe J, Clair AH, Bally JF, Riquier F, et al. Therapies for obsessive-compulsive disorder: Current state of the art and perspectives for approaching treatment-resistant patients. Front Psychiatry. 2023;14:1065812.

3. Abramowitz JS, Blakey SM, Reuman L, Buchholz JL. New Directions in the Cognitive-Behavioral Treatment of OCD: Theory, Research, and Practice. Behavior Therapy. 2018;49(3):311–22.

4. Spencer SD, Stiede JT, Wiese AD, Goodman WK, Guzick AG, Storch EA. Cognitive-Behavioral Therapy for Obsessive-Compulsive Disorder. Psychiatr Clin North Am. 2023;46(1):167–80.

5. Öst LG, Enebrink P, Finnes A, Ghaderi A, Havnen A, Kvale G, et al. Cognitive behavior therapy for obsessive-compulsive disorder in routine clinical care: A systematic review and meta-analysis. Behav Res Ther. 2022;159:104170.

6. O’Neill, J., & Feusner, J. D. (2015). Cognitive-behavioral therapy for obsessive-compulsive disorder: access to treatment, prediction of long-term outcome with neuroimaging. Psychol Res Behav Manag, 8, 211–223. 10.2147/prbm.s75106

7. Mukai K, Kyosuke Y, Ogino S, Hosoi Y, Hayashida K, Matsunaga H. Benefits and barriers associated with using cognitive-behavioral therapy to treat obsessive-compulsive disorder: a narrative review. Front Psychiatry. 2025;16:1593384.

8. Olatunji BO, Davis ML, Powers MB, Smits JAJ. Cognitive-behavioral therapy for obsessive-compulsive disorder: A meta-analysis of treatment outcome and moderators. Journal of Psychiatric Research. 2013;47(1):33–41.

9. Knopp J, Knowles S, Bee P, Lovell K, Bower P. A systematic review of predictors and moderators of response to psychological therapies in OCD: Do we have enough empirical evidence to target treatment? Clinical Psychology Review. 2013;33(8):1067–81.

10. Keeley ML, Storch EA, Merlo LJ, Geffken GR. Clinical predictors of response to cognitive-behavioral therapy for obsessive–compulsive disorder. Clinical Psychology Review. 2008;28(1):118–30.

11. Machado-Sousa M, Bertolín S, Picó-Pérez M, Costa AD, Vieira R, Alonso P, et al. Neurobiological correlates of CBT response in OCD through the analysis of resting state networks. International Journal of Clinical and Health Psychology. 2025;25(2).

12. Li P, Yang X, Greenshaw AJ, Li S, Luo J, Han H, et al. The effects of cognitive behavioral therapy on resting-state functional brain network in drug-naive patients with obsessive–compulsive disorder. Brain and Behavior. 2018;8(5):e00963.

13. Reggente N, Moody TD, Morfini F, Sheen C, Rissman J, O’Neill J, et al. Multivariate resting-state functional connectivity predicts response to cognitive behavioral therapy in obsessive-compulsive disorder. Proc Natl Acad Sci U S A. 2018;115(9):2222–7.

14. van de Mortel LA, Bruin WB, Alonso P, Bertolín S, Feusner JD, Guo J, et al. Development and validation of a machine learning model to predict cognitive behavioral therapy outcome in obsessive-compulsive disorder using clinical and neuroimaging data. Journal of Affective Disorders. 2025;389:119729.

15. Thai M, Olson EA, Nickels S, Dillon DG, Webb CA, Ren B, et al. Neural and behavioral markers of inhibitory control predict symptom improvement during internet-delivered cognitive behavioral therapy for depression. Translational Psychiatry. 2024;14(1):303.

16. Falconer E, Allen A, Felmingham KL, Williams LM, Bryant RA. Inhibitory neural activity predicts response to cognitive-behavioral therapy for posttraumatic stress disorder. J Clin Psychiatry. 2013;74(9):895–901.

17. Norman LJ, Mannella KA, Yang H, Angstadt M, Abelson JL, Himle JA, et al. Treatment-Specific Associations Between Brain Activation and Symptom Reduction in OCD Following CBT: A Randomized fMRI Trial. Am J Psychiatry. 2021;178(1):39–47.

18. Postma TS, Fitzsimmons S, Vriend C, Batelaan NM, van der Werf YD, van den Heuvel OA. Transcranial Magnetic Stimulation-Induced Plasticity Improving Cognitive Control in Obsessive-Compulsive Disorder, Part II: Task-Based Neural Predictors of Treatment Response. Biol Psychiatry. 2024.

19. Grützmann R, Klawohn J, Elsner B, Reuter B, Kaufmann C, Riesel A, et al. Error-related activity of the sensorimotor network contributes to the prediction of response to cognitive-behavioral therapy in obsessive–compulsive disorder. NeuroImage: Clinical. 2022;36:103216.

20. Goodman KW. The Yale-Brown Obsessive Compulsive Scale. Archives of General Psychiatry. 1989;46(11):1006.

21. Scahill L, Riddle AM, Mcswiggin-Hardin M, Ort IS, King AR, Goodman KW, et al. Children’s Yale-Brown Obsessive Compulsive Scale: Reliability and Validity. Journal of the American Academy of Child & Adolescent Psychiatry. 1997;36(6):844–52.

22. Dzinalija, N., van den Heuvel, O. A., Simpson, H. B., Ivanov, I., Araujo, A., Balachander, S.,…Veer, I. M. (2025). Inhibitory control and error processing in Obsessive-Compulsive Disorder: A mega-analysis of task-based fMRI data by the ENIGMA-OCD consortium. bioRxiv, 2025.2010.2022.683868. 10.1101/2025.10.22.683868

23. Farris SG, McLean CP, Van Meter PE, Simpson HB, Foa EB. Treatment response, symptom remission, and wellness in obsessive-compulsive disorder. J Clin Psychiatry. 2013;74(7):685–90.

24. Waller L, Erk S, Pozzi E, Toenders YJ, Haswell CC, Büttner M, et al. ENIGMA HALFpipe: Interactive, reproducible, and efficient analysis for resting-state and task-based fMRI data. Hum Brain Mapp. 2022;43(9):2727–42.

25. Norman LJ, Taylor SF, Liu Y, Radua J, Chye Y, De Wit SJ, et al. Error Processing and Inhibitory Control in Obsessive-Compulsive Disorder: A Meta-analysis Using Statistical Parametric Maps. Biol Psychiatry. 2019;85(9):713–25.

26. Schaefer A, Kong R, Gordon ME, Laumann OT, Zuo X-N, Holmes JA, et al. Local-Global Parcellation of the Human Cerebral Cortex from Intrinsic Functional Connectivity MRI. Cerebral Cortex. 2018;28(9):3095–114.

27. Tian Y, Margulies SD, Breakspear M, Zalesky A. Topographic organization of the human subcortex unveiled with functional connectivity gradients. Nature Neuroscience. 2020;23(11):1421–32.

28. Johnson WE, Li C, Rabinovic A. Adjusting batch effects in microarray expression data using empirical Bayes methods. Biostatistics. 2007;8(1):118–27.

29. R Core Team. R: A Language and Environment for Statistical Computing. Vienna, Austria: R Foundation for Statistical Computing; 2024.

30. Chen G, Xiao Y, Taylor PA, Rajendra JK, Riggins T, Geng F, et al. Handling Multiplicity in Neuroimaging Through Bayesian Lenses with Multilevel Modeling. Neuroinformatics. 2019;17(4):515–45.

31. Gildenblat J. A python library for confidence intervals. GitHub; 2023.

32. Pagliaccio D, Middleton R, Hezel D, Steinman S, Snorrason I, Gershkovich M, et al. Task-based fMRI predicts response and remission to exposure therapy in obsessive-compulsive disorder. Proc Natl Acad Sci U S A. 20190923 ed 2019. p. 20346–53.

33. Leonards CA, Harrison BJ, Jamieson AJ, Steward T, Lux S, Philipsen A, et al. A distinct intra-individual suppression subnetwork in the brain’s default mode network across cognitive tasks. Cereb Cortex. 2023;33(8):4553–61.

34. Olatunji BO, Ferreira-Garcia R, Caseras X, Fullana MA, Wooderson S, Speckens A, et al. Predicting response to cognitive behavioral therapy in contamination-based obsessive-compulsive disorder from functional magnetic resonance imaging. Psychol Med. 2014;44(10):2125–37.

35. Gonçalves ÓF, Soares JM, Carvalho S, Leite J, Ganho-Ávila A, Fernandes-Gonçalves A, et al. Patterns of Default Mode Network Deactivation in Obsessive Compulsive Disorder. Scientific Reports. 2017;7(1):44468.

36. Zhao XH, Wang PJ, Li CB, Hu ZH, Xi Q, Wu WY, et al. Altered default mode network activity in patient with anxiety disorders: an fMRI study. Eur J Radiol. 2007;63(3):373–8.

37. Lucherini Angeletti L, Scalabrini A, Ricca V, Northoff G. Topography of the Anxious Self: Abnormal Rest-Task Modulation in Social Anxiety Disorder. Neuroscientist. 2023;29(2):221–44.

38. Yuan S, Wu H, Wu Y, Xu H, Yu J, Zhong Y, et al. Neural Effects of Cognitive Behavioral Therapy in Psychiatric Disorders: A Systematic Review and Activation Likelihood Estimation Meta-Analysis. Front Psychol. 2022;13:853804.

39. Moody TD, Morfini F, Cheng G, Sheen C, Tadayonnejad R, Reggente N, et al. Mechanisms of cognitive-behavioral therapy for obsessive-compulsive disorder involve robust and extensive increases in brain network connectivity. Transl Psychiatry. 2017;7(9):e1230.

40. Bruin W, Denys D, van Wingen G. Diagnostic neuroimaging markers of obsessive-compulsive disorder: Initial evidence from structural and functional MRI studies. Prog Neuropsychopharmacol Biol Psychiatry. 2019;91:49–59.

41. van Velzen, L. S., Vriend, C., de Wit, S. J., & van den Heuvel, O. A. (2014). Response inhibition and interference control in obsessive-compulsive spectrum disorders. Frontiers in human neuroscience, 8, 419. 10.3389/fnhum.2014.00419

42. Picó-Pérez M, Fullana MA, Albajes-Eizagirre A, Vega D, Marco-Pallarés J, Vilar A, et al. Neural predictors of cognitive-behavior therapy outcome in anxiety-related disorders: a meta-analysis of task-based fMRI studies. Psychological Medicine. 2023;53(8):3387–95.

43. Hahn T, Kircher T, Straube B, Wittchen H-U, Konrad C, Ströhle A, et al. Predicting Treatment Response to Cognitive Behavioral Therapy in Panic Disorder With Agoraphobia by Integrating Local Neural Information. JAMA Psychiatry. 2015;72(1):68–74.

44. Queirazza F, Fouragnan E, Steele JD, Cavanagh J, Philiastides MG. Neural correlates of weighted reward prediction error during reinforcement learning classify response to cognitive behavioral therapy in depression. Sci Adv. 2019;5(7):eaav4962.

45. Costafreda SG, Khanna A, Mourao-Miranda J, Fu CHY. Neural correlates of sad faces predict clinical remission to cognitive behavioural therapy in depression. NeuroReport. 2009;20(7).

46. Ball TM, Stein MB, Ramsawh HJ, Campbell-Sills L, Paulus MP. Single-Subject Anxiety Treatment Outcome Prediction using Functional Neuroimaging. Neuropsychopharmacology. 2014;39(5):1254–61.

47. Frick A, Engman J, Alaie I, Björkstrand J, Gingnell M, Larsson E-M, et al. Neuroimaging, genetic, clinical, and demographic predictors of treatment response in patients with social anxiety disorder. Journal of Affective Disorders. 2020;261:230–7.

48. Schnack HG, Kahn RS. Detecting Neuroimaging Biomarkers for Psychiatric Disorders: Sample Size Matters. Front Psychiatry. 2016;7:50.

49. Hilbert K, Böhnlein J, Meinke C, Chavanne AV, Langhammer T, Stumpe L, et al. Lack of evidence for predictive utility from resting state fMRI data for individual exposure-based cognitive behavioral therapy outcomes: A machine learning study in two large multi-site samples in anxiety disorders. Neuroimage. 2024;295:120639.

50. Harris JK, Hassel S, Davis AD, Zamyadi M, Arnott SR, Milev R, et al. Predicting escitalopram treatment response from pre-treatment and early response resting state fMRI in a multi-site sample: A CAN-BIND-1 report. NeuroImage: Clinical. 2022;35:103120.

51. Poldrack RA, Huckins G, Varoquaux G. Establishment of Best Practices for Evidence for Prediction: A Review. JAMA Psychiatry. 2020;77(5):534–40.

52. Bzdok D, Varoquaux G, Steyerberg EW. Prediction, Not Association, Paves the Road to Precision Medicine. JAMA Psychiatry. 2021;78(2):127–8.

53. Huyser C, Veltman DJ, Wolters LH, de Haan E, Boer F. Developmental aspects of error and high-conflict-related brain activity in pediatric obsessive–compulsive disorder: a fMRI study with a Flanker task before and after CBT. Journal of Child Psychology and Psychiatry. 2011;52(12):1251–60.

54. van der Straten A, Bruin W, van de Mortel L, ten Doesschate F, Merkx MJM, de Koning P, et al. Pharmacological and Psychological Treatment Have Common and Specific Effects on Brain Activity in Obsessive-Compulsive Disorder. Depression and Anxiety. 2024;2024(1):6687657.

55. Hermida-Barros L, García-Delgar B, Lera-Miguel S, Forcadell E, Moreno E, Primé-Tous M, et al. Concentrated Cognitive-Behavior Therapy for Unmedicated Children and Adolescents With Obsessive-Compulsive Disorder in Routine Clinical Care: A Randomized Controlled Trial With a 6-Month Naturalistic Follow-up. Behavior Therapy. 2025;56(4):799–811.

56. Machado-Sousa, M., Bertolín, S., Picó-Pérez, M., Costa, A. D., Vieira, R., Alonso, P.,…Soriano-Mas, C. (2025). Neurobiological correlates of CBT response in OCD through the analysis of resting state networks. Int J Clin Health Psychol, 25(2), 100585. 10.1016/j.ijchp.2025.100585

57. Lillevik Thorsen A, de Wit SJ, Hagland P, Ousdal OT, Hansen B, Hagen K, et al. Stable inhibition-related inferior frontal hypoactivation and fronto-limbic hyperconnectivity in obsessive-compulsive disorder after concentrated exposure therapy. Neuroimage Clin. 2020;28:102460.

