## Supplement for "Predicting cognitive-behavioral therapy outcomes in obsessive-compulsive disorder from inhibitory control neural activity: A mega-analysis and machine learning study from the ENIGMA-OCD consortium"

Table S1 – Demographics of included participants from each sample. Values denote mean (SD) unless otherwise stated. (C)Y-BOCS = (Children’s) Yale-Brown Obsessive-Compulsive Scale.

| **PI, Sample** | **n** | **% Male** | **Age** | **% Child onset** | **% Medicated** | **(C)Y-BOCS pre CBT** | **(C)Y-BOCS post CBT** | **% Responders** | **% Remitters** |
| --- | --- | --- | --- | --- | --- | --- | --- | --- | --- |
| Huyser,  Amsterdam | 27 | 37.04 | 13.19 ± 2.57 | 100 | 0 | 24.78 ± 5.13 | 13.41 ± 9.3 | 66.67 | 44.44 |
| van Wingen, Amsterdam | 14 | 64.29 | 30.64 ± 9.39 | 57.14 | 7.14 | 25 ± 3.14 | 12.29 ± 6.26 | 71.43 | 42.86 |
| Fullana,  Barcelona | 31 | 58.06 | 16.87 ± 8.49 | 93.55 | 0 | 25.03 ± 4.5 | 15.19 ± 7.48 | 67.74 | 41.94 |
| Menchon/  Soriano-Mas, Barcelona | 22 | 45.45 | 35.59 ± 9.94 | 59.09 | 100 | 24.86 ± 6.47 | 18.09 ± 6.92 | 40.91 | 27.27 |
| Thorsen,  Bergen | 36 | 36.11 | 30.36 ± 8.69 | 41.67 | 25 | 26.83 ± 4.42 | 11.69 ± 6.46 | 83.33 | 55.56 |

### Table S2 – Details of Cognitive Behavioral Therapy (CBT) administered to each sample.

| **PI, Sample** | **Duration of**  **CBT (hours)** | **Duration of**  **CBT (weeks)** | **Exposure with response prevention** | **Individual / group sessions** | **Homework tasks** |
| --- | --- | --- | --- | --- | --- |
| Huyser, Amsterdam | 16 | 16 | Yes | Individual | Yes |
| van Wingen, Amsterdam | 16 | 16 | Yes | Mix (n=13 group,  n=4 individual) | Yes |
| Fullana, Barcelona | 20 | 4 | Yes | Individual | Yes |
| Menchon/Soriano-Mas, Barcelona | 16 | 11-27 (mean=20.3±4.0) | Yes | Group | Yes |
| Thorsen, Bergen | 24 | 1 | Yes | Group | Yes |

### Table S3 – MRI acquisition parameters of each sample

| **PI, Sample** | **Scanner vendor**  **and type** | **Head coil**  **(N channels)** | **Pulse  sequence** | **Single-/ multiband** | **Matrix** | **Volumes  (N)** | **Slices  (N)** | **Scan order** | **Slicegap  (mm)** | **TR  (ms)** | **TE  (ms)** | **Flipangle  (°)** | **FOV (mm)** | **Voxel size  (mm)** |
| --- | --- | --- | --- | --- | --- | --- | --- | --- | --- | --- | --- | --- | --- | --- |
| Huyser, Amsterdam | 3T Philip Intera MR | 6 | GE-EPI | singleband | 96x96 | 250 | 40 | interleaved | 0 | 2300 | 30 | 90 | 220x120x220 | 2.29x  2.29x3 |
| van Wingen, Amsterdam | 3T Philips Achieva | 32 | GE-EPI | singleband | 75x75 | 197 | 37 | interleaved | 0.3 | 2375 | 26 | 76 | 224x224 | 2.8x  2.8x3 |
| Fullana, Barcelona | 3T Philips Ingenia | 32 | GE-EPI | singleband | 96x96 | 100 | 32 | interleaved | 0.75 | 2000 | 30 | 70 | 240x240 | 2.5x2.5x4.25 |
| Menchon/  Soriano-Mas, Barcelona | 3T Philips Ingenia | 32 | SS-EPI | singleband | 80x80 | 97 | 40 | interleaved | 0 | 2000 | 25 | 90 | 240 | 3mm3 |
| Thorsen, Bergen | 3T GE Discovery MR750 | 8 | GE-EPI | singleband | 64x64 | 430 | 34 | interleaved | 0.2 | 2100 | 30 | 80 | 220 | 3.44x  3.44x3 |

Table S4 - Regions of interest Identified from Norman et al. (2019;(11)) showing whole-brain activity in healthy controls during inhibitory control tasks for the response inhibition and error processing contrasts. Coordinates are in MNI152 NLIN 6th generation space. ACC = anterior cingulate cortex; aI/fO = anterior insula/frontal operculum; dlPFC = dorsolateral prefrontal cortex; Occ = occipital; PMC = primary motor cortex; SMA = supplementary motor area; SMG = supramarginal gyrus; SPL = superior parietal lobule; STG = superior temporal gyrus.

| **Region** | **Lateralization** | **MNI coordinates** | | |
| --- | --- | --- | --- | --- |
|  |  | **x** | **y** | **z** |
| **Response inhibition [stop success > go]** | | | | |
| aI/fO | right | 34 | 18 | 8 |
|  | left | -40 | 10 | 2 |
| SMA | right | 4 | 14 | 58 |
|  | left | -4 | 14 | 58 |
| SPL | right | 42 | -44 | 58 |
|  | left | -42 | -44 | 58 |
| Occ. lobe | right | 28 | -72 | 36 |
|  | left | -28 | -72 | 36 |
| PMC | right | 38 | -8 | 62 |
|  | left | -48 | 2 | 24 |
| dlPFC | right | 34 | 44 | 34 |
|  | left | -34 | 44 | 34 |
| **Error processing [stop fail > stop success]** | | | | |
| aI/fO | right | 44 | 14 | 2 |
|  | left | -48 | 16 | -2 |
| STG | left | -40 | -8 | -12 |
| ACC | bilateral | 0 | 34 | 32 |
| Occ. lobe | bilateral | 0 | -82 | 18 |
| SMG | right | 66 | -40 | 28 |
|  | left | -58 | -46 | 30 |

**Response inhibition	Error processing**
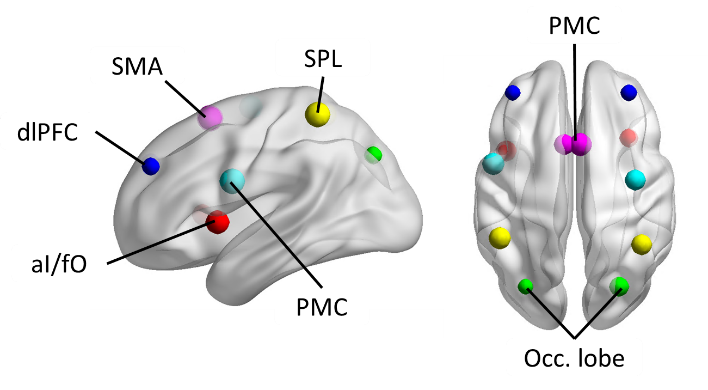

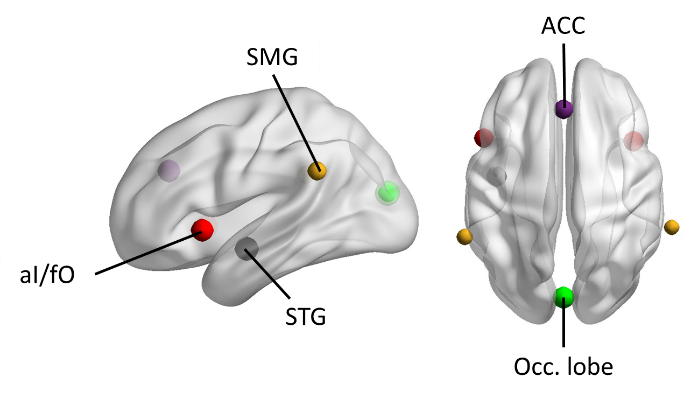

### Preprocessing and quality control guidelines

Preprocessing was done at each site according to harmonized guidelines and each site performed quality-control on their own preprocessed data following harmonized guidelines which can be retrieved from the online repository [doi.org/10.5281/zenodo.17434990](https://doi.org/10.5281/zenodo.17434990).

### Missing brain activity data

Brain activity data were included in ROI or whole-brain parcel analyses only if at least 30% of voxels in the ROI or parcel contained signal. ROIs or parcels not meeting this threshold were excluded; missing values were not imputed. No ROIs failed this criterion for either response inhibition or error processing contrasts. In whole-brain analyses, some parcels near the edges of the brain had insufficient coverage across contrasts.

**Table S5 – Whole-brain parcels missing data per analysis, ordered by percent of sample missing data in given parcel** L = left hemisphere; OFC = orbitofrontal cortex; PFC = prefrontal cortex; Post = posterior; R = right hemisphere; Temp = temporal.

| **Parcel** | **% missing** |
| --- | --- |
| **Response inhibition** | |
| Limbic OFC 1 R | 38.5 |
| Limbic OFC 2 L | 30.0 |
| Limbic TempPole 2 R | 20.0 |
| Limbic TempPole 1 L | 16.9 |
| Limbic TempPole 1 R | 12.3 |
| Default Temp 2 L | 9.2 |
| Limbic TempPole 2 L | 9.2 |
| Limbic TempPole 3 R | 5.4 |
| Limbic TempPole 3 L | 3.8 |
| Visual 5 L | 3.1 |
| Control Temp 1 R | 0.8 |
| Default PFC 13 L | 0.8 |
| Default Temp 1 L | 0.8 |
| Default Temp 2 R | 0.8 |
| Dorsal Attention Post 10 L | 0.8 |
| Dorsal Attention Post 10 R | 0.8 |
| **Error processing** | |
| Limbic OFC 1 R | 38.5 |
| Limbic OFC 2 L | 30.0 |
| Limbic TempPole 2 R | 20.0 |
| Limbic TempPole 1 L | 16.9 |
| Limbic TempPole 1 R | 12.3 |
| Default Temp 2 L | 9.2 |
| Limbic TempPole 2 L | 9.2 |
| Limbic TempPole 3 R | 5.4 |
| Limbic TempPole 3 L | 3.8 |
| Visual 5 L | 3.1 |
| Control Temp 1 R | 0.8 |
| Default PFC 13 L | 0.8 |
| Default Temp 1 L | 0.8 |
| Default Temp 2 R | 0.8 |
| Dorsal Attention Post 10 L | 0.8 |
| Dorsal Attention Post 10 R | 0.8 |

Table S6 – Results of univariate analyses Only regions that show significant results after false-discovery rate (FDR) correction are displayed. Regions of interest (ROIs) refer to predefined spheres around coordinates reported in Norman et al. (7) for response inhibition and error processing. Whole-brain regions refer to the Schaefer 200-parcel 7-network cortical atlas and Melbourne 32-region subcortical atlas. Δ (C)Y-BOCS = change in (Children’s) Yale-Brown Obsessive-Compulsive Scale total score (pre-treatment minus post-treatment); DA = dorsal anterior; DF = degrees of freedom; L = left; pCunPCC = precuneus/posterior cingulate cortex; PFC = prefrontal cortex; PFCl = lateral prefrontal cortex; PFCmp = medial prefrontal cortex; PFCdPFCm = dorsolateral/medial prefrontal cortex; PMC = primary motor cortex; R = right; ROI = region of interest; SMA = supplementary motor area; SMG = supramarginal gyrus; SPL = superior parietal lobule; VA = ventral anterior.

| **Features** | **Outcome** | **Region** | **T value** | **DF** | **P_FDR corr._** | **Cohen's *d*** |
| --- | --- | --- | --- | --- | --- | --- |
| **Response Inhibition** | | | | | | |
| ROIs | Δ (C)Y-BOCS | SPL (R) | -3.484 | 125 | 0.008 | -0.611 |
|  |  | PMC (R) | -2.935 | 125 | 0.016 | -0.515 |
|  |  | SMA (R) | -2.950 | 125 | 0.016 | -0.517 |
|  |  | SMA (L) | -2.705 | 125 | 0.023 | -0.474 |
| Whole-brain parcels | Δ (C)Y-BOCS | Default PFC 1 (L) | -3.663 | 125 | 0.028 | -0.643 |
|  |  | Default pCunPCC 3 (L) | -3.672 | 125 | 0.028 | -0.644 |
|  |  | Posterior Hippocampus (R) | -3.678 | 125 | 0.028 | -0.645 |
|  |  | Default PFC 5 (L) | -3.179 | 125 | 0.034 | -0.558 |
|  |  | Visual 9 (L) | -3.168 | 125 | 0.034 | -0.556 |
|  |  | Control Cingulate 1 (R) | -3.247 | 125 | 0.034 | -0.570 |
|  |  | Control PFCmp 2 (R) | -3.242 | 125 | 0.034 | -0.569 |
|  |  | Control Temporal 1 (R) | -3.256 | 125 | 0.034 | -0.571 |
|  |  | Default PFCdPFCm 3 (R) | -3.239 | 125 | 0.034 | -0.568 |
|  |  | Default pCunPCC 2 (R) | -3.431 | 125 | 0.034 | -0.602 |
|  |  | Default pCunPCC 3 (R) | -3.191 | 125 | 0.034 | -0.560 |
|  |  | Thalamus VA (L) | -3.378 | 125 | 0.034 | -0.593 |
|  |  | Posterior Hippocampus (L) | -3.402 | 125 | 0.034 | -0.597 |
|  |  | Dorsal Attention Posterior 7 (R) | -3.137 | 125 | 0.035 | -0.550 |
|  |  | Default PFC 11 (L) | -3.069 | 125 | 0.038 | -0.538 |
|  |  | Control PFCl 7 (R) | -3.046 | 125 | 0.038 | -0.534 |
|  |  | Default PFCdPFCm 2 (R) | -3.033 | 125 | 0.038 | -0.532 |
|  |  | Dorsal Attention Posterior 6 (R) | -3.080 | 125 | 0.038 | -0.540 |
|  |  | Control Parietal 3 (R) | -2.985 | 125 | 0.041 | -0.524 |
|  |  | Thalamus DA (R) | -2.974 | 125 | 0.041 | -0.522 |
|  |  | Visual 5 (R) | -2.926 | 125 | 0.045 | -0.513 |
|  |  | Control Cingulate 1 (L) | -2.910 | 125 | 0.045 | -0.510 |
|  |  | Control PFCl 4 (L) | -2.806 | 125 | 0.048 | -0.492 |
|  |  | Default PFC 4 (L) | -2.805 | 125 | 0.048 | -0.492 |
|  |  | Visual 3 (L) | -2.810 | 125 | 0.048 | -0.493 |
|  |  | Control Cingulate 2 (R) | -2.798 | 125 | 0.048 | -0.491 |
|  |  | Default Parietal 2 (R) | -2.829 | 125 | 0.048 | -0.496 |
|  |  | Default Parietal 3 (R) | -2.831 | 125 | 0.048 | -0.497 |
|  |  | Visual 2 (R) | -2.859 | 125 | 0.048 | -0.502 |
|  |  | Dorsal Attention Posterior 4 (R) | -2.771 | 125 | 0.050 | -0.486 |
| Whole-brain parcels | Remission | Default pCunPCC 3 (L) | -3.784 | 126 | 0.028 | -0.664 |
|  |  | Default pCunPCC 2 (R) | -3.826 | 126 | 0.028 | -0.671 |
| **Error Processing** | | | | | | |
| ROIs | Remission | SMG (R) | -2.756 | 126 | 0.047 | -0.483 |
| Whole-brain parcels | Δ (C)Y-BOCS | Default pCunPCC 2 (L) | 3.855 | 125 | 0.040 | 0.676 |
|  |  | Default pCunPCC 2 (R) | 3.677 | 125 | 0.040 | 0.645 |

### Sensitivity Analyses

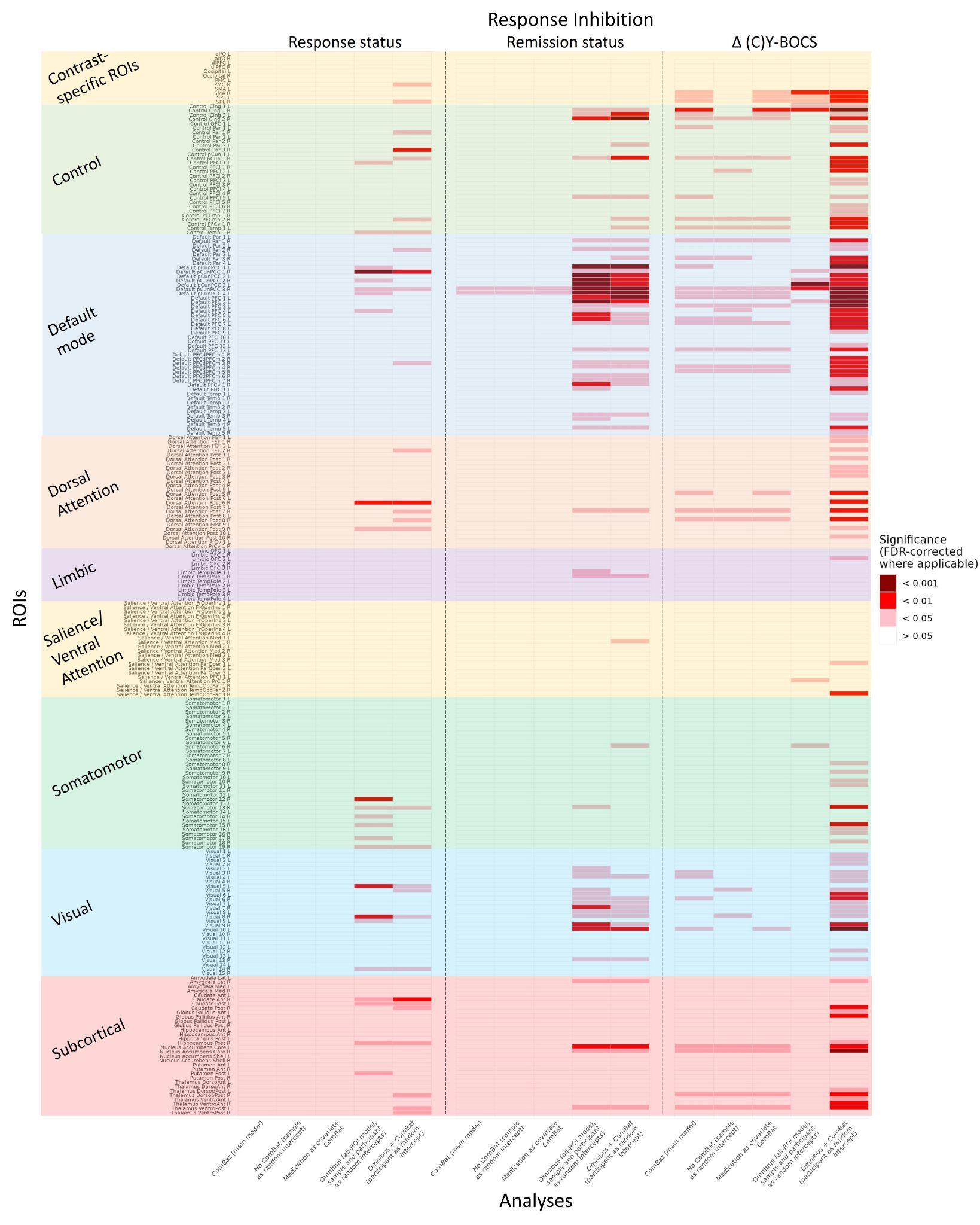

**Figure S1 – Sensitivity analyses using alternative mixed-effects models for response inhibition** Displayed are p-values for each ROI or whole-brain parcel, testing associations with response status, remission status, and change in symptom severity from pre- to post-CBT treatment. Models included: 1) ComBat harmonization of samples (main model reported in the manuscript); 2) random intercept for sample, 3) ComBat-harmonized model additionally controlling for medication status; 4) simultaneous modelling of all ROIs with random intercepts for participant and sample, and 5) ComBat-harmonized simultaneous modelling of all ROIs (participant as a random intercept). P-values were FDR-corrected in models that fit one ROI at a time. aI/fO = anterior insula/frontal operculum; Ant = anterior; Cing = cingulate; Δ (C)Y-BOCS = change in (Children’s) Yale-Brown Obsessive-Compulsive Scale total score (pre-treatment minus post-treatment); dlPFC = dorsolateral prefrontal cortex; DorsoAnt = dorsal anterior; DorsopPost = dorsoposterior; FEF = frontal eye fields; FrOperIns = frontal operculum / insula; L = left; Lat = lateral; Med = medial; Occ = occipital; OFC = orbitofrontal cortex; Par = parietal; ParOper = parietal operculum; PCC = posterior cingulate cortex; pCun = precuneus; PFCl = lateral prefrontal cortex; PFC = prefrontal cortex; PFCdPFCm = dorsal/medial prefrontal cortex; PFCmp = medial–posterior prefrontal cortex; PFCv = ventral prefrontal cortex; PHC = parahippocampal cortex; PMC = primary motor cortex; Post = posterior; PrC = precentral; PrCv = precentral ventral; R = right; SMA = supplementary motor area; SPL = superior parietal lobule; Temp = temporal; TempOccPar = temporo-occipito-parietal; VentroAnt = ventroanterior; VentroPost = ventroposterior.

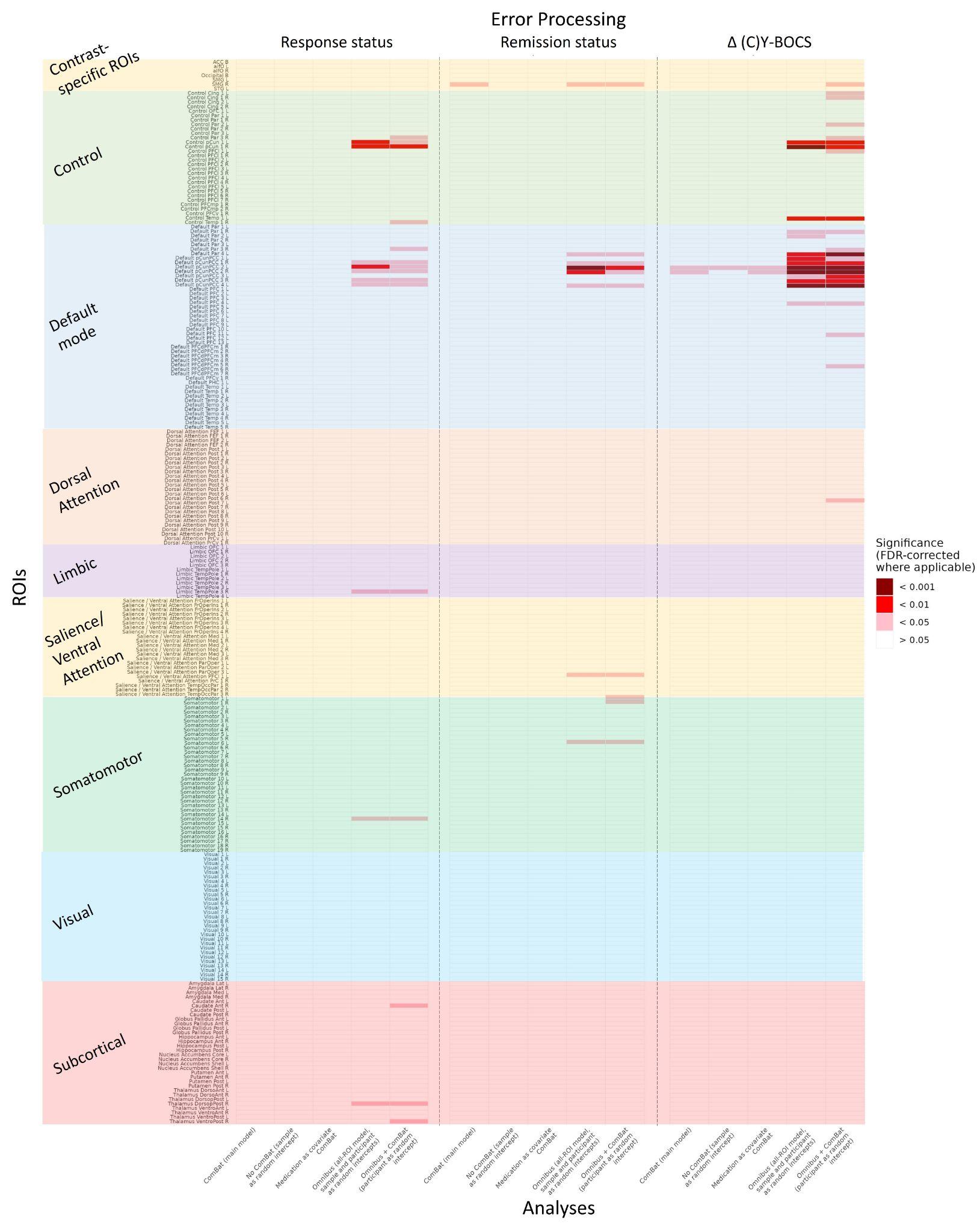

**Figure S2 – Sensitivity analyses using alternative mixed-effects models for error processing** ACC = anterior cingulate cortex; aI/fO = anterior insula/frontal operculum; Ant = anterior; Cing = cingulate; Δ (C)Y-BOCS = change in (Children’s) Yale-Brown Obsessive-Compulsive Scale total score (pre-treatment minus post-treatment); DorsopPost = dorsoposterior; DorsoAnt = dorsal anterior; FEF = frontal eye fields; FrOperIns = frontal operculum / insula; L = left; Lat = lateral; Med = medial; OFC = orbitofrontal cortex; Occ = occipital; PCC = posterior cingulate cortex; PFCl = lateral prefrontal cortex; PFC = prefrontal cortex; PFCdPFCm = dorsal/medial prefrontal cortex; PFCmp = medial–posterior prefrontal cortex; PFCv = ventral prefrontal cortex; PHC = parahippocampal cortex; Par = parietal; ParOper = parietal operculum; Post = posterior; PrC = precentral; PrCv = precentral ventral; R = right; SMG = supramarginal gyrus; STG = superior temporal gyrus; Temp = temporal; TempOccPar = temporo-occipito-parietal; VentroAnt = ventroanterior; VentroPost = ventroposterior.

**Tables S7 – Full output of univariate analyses and sensitivity analyses**

The full output of the univariate ROI and whole-brain analyses presented in Table S6 of the manuscript can be retrieved from the online repository [doi.org/10.5281/zenodo.17434990](https://doi.org/10.5281/zenodo.17434990). Additionally, full output is available from sensitivity analyses with alternate univariate mixed-effects models including a random intercept for sample, combined mixed-effects models including all ROIs simultaneously (with random intercepts for sample and participant), and models including medication status as a covariate. Finally, ridge plots of whole-brain Bayesian multilevel models presented in Figures S3-7 of the supplement are available.

### Bayesian analyses

Each participant’s ROI and whole-brain activity values were entered into group analyses using a Bayesian multilevel model (RBA, v1.0.10;(12)). This approach simultaneously analyses all regions, considering the shared non-independent information across brain regions within a participant in one statistical model. Unlike null hypothesis significance testing, this approach allows us to directly assess the credibility of our hypothesis. A detailed discussion of this approach is provided in Dzinalija et al. (13) and Chen et al. (12). We used a noninformative Gaussian prior and four Markov chains with 4,000 permutations per chain, confirming convergence by ensuring Rhat <1.1. The credibility of an effect was inferred from the area under the curve of the positive posterior distribution, summarized as the positive posterior probability (P+). We interpreted P+ values as follows: <0.10 or >0.90 indicated moderate evidence of an effect, <0.05 or >0.95 indicated strong evidence, and <0.025 or >0.975 indicated very strong evidence (12).

Analyses were conducted separately for each contrast (response inhibition and error processing) on each outcome measure: treatment response, remission status, and change in symptom severity from pre- to post-CBT. We assessed the main effect of response inhibition and error processing across all tasks and all participants using an intercept model. We additionally conducted 1) pairwise comparisons of responders vs. non-responders to CBT; 2) pairwise comparisons of remitters vs. non-remitters; and 3) continuous analyses of pre-to-post-CBT change in (C)Y-BOCS scores. Sample, age, and sex were included as covariates of no interest in all analyses. We additionally re-ran all analyses after performing ComBat harmonization on the dataset.

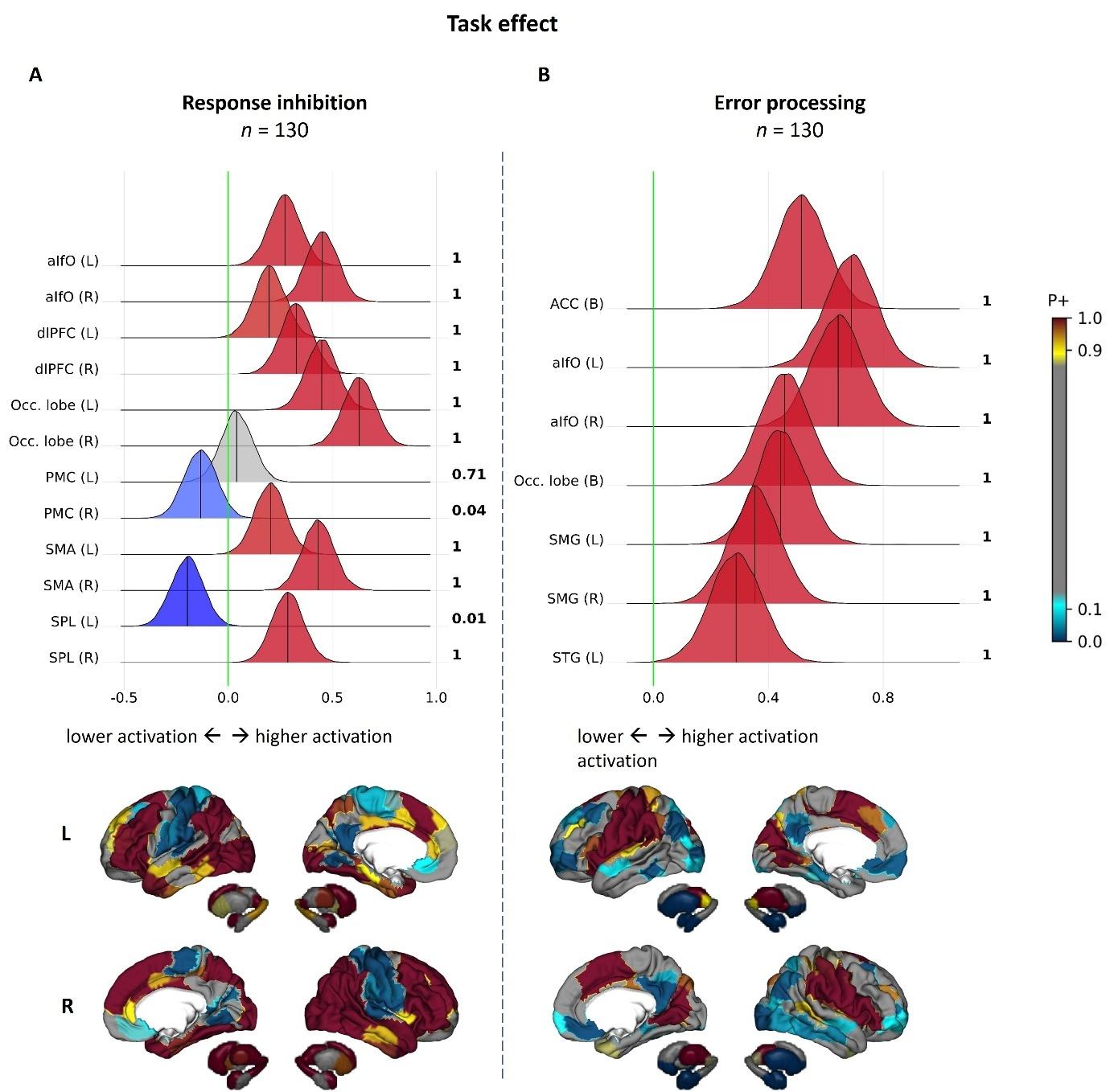

**Figure S3 – Main effect of response inhibition and error processing across all participants** Results from Bayesian multilevel analyses of group-level contrasts for (A) response inhibition and (B) error processing across all tasks. For each contrast, the top panels display region-of-interest results, while the bottom panel displays whole-brain results. In the ROI panels, posterior probability distributions express the credibility of an effect in each region. Next to each distribution the posterior probability of a positive effect (P+) is shown in bold. Distributions to the right of the green no-effect line indicate stronger activity, while those to the left indicate weaker activity. Regions are color-coded to reflect the strength of evidence: (darker) red = stronger evidence for activity (P+ values > 0.90 indicate moderate to very strong evidence for a positive effect), (darker) blue = stronger evidence for deactivity (P+ values < 0.10 indicate moderate to very strong evidence for a negative effect), grey = no strong evidence of activity or deactivity. Values on the x-axis represent the difference in regional activity levels between experimental and baseline conditions for each contrast (expressed as difference in Z-scores). In the whole-brain panels, P+ values denote the probability of stronger brain activity in a given region of the Schaefer 200-parcel 7-network cortical atlas and Melbourne 32-region subcortical atlas. Displayed are lateral and medial views of the cortex and subcortex. ACC = anterior cingulate cortex; aI/fO = anterior insula/frontal operculum; dlPFC = dorsolateral prefrontal cortex; L = left;  Occ = occipital; PMC = primary motor cortex; R = right; SMA = supplementary motor area; SMG = supramarginal gyrus; SPL = superior parietal lobule; STG = superior temporal gyrus.

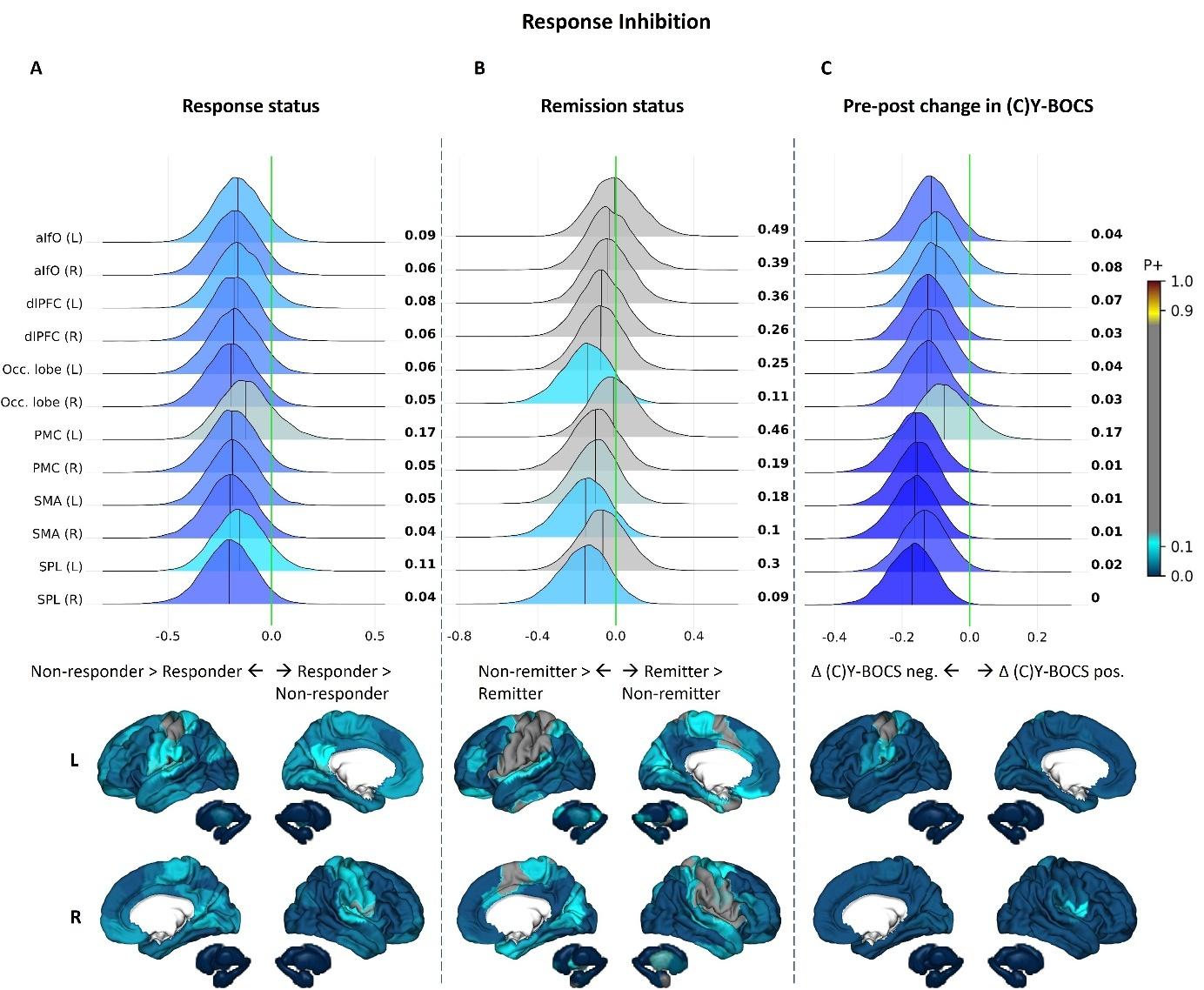

**Figure S4 – Effect of CBT outcomes on response inhibition** Results from Bayesian multilevel analyses of the relation between pre-treatment brain activity during response inhibition and (A) response to CBT, (B) remission after CBT, and (C) pre-to-post-CBT change in symptom severity. Sample effects were corrected for by including sample as a covariate of no interest. For each outcome measure, region-of-interest results are shown in the top panels and whole-brain results in the bottom panels. In the ROI plots, distributions to the right of the no-effect line indicate stronger activity in responders/remitters relative to (A,B) non-responders/non-remitters, or (C) a positive association between activity and change in (C)Y-BOCS scores. Distributions to the left of the no-effect line indicate stronger activity in (A,B) non-responders/non-remitters, or (C) a negative association with symptom change. Values on the x-axis represent, for categorical outcomes (A,B), the difference in regional activity between groups (expressed as difference in Z-scores), and for the continuous outcome (C), the change in activity associated with greater pre-to-post-treatment change in (C)Y-BOCS scores. aI/fO = anterior insula/frontal operculum; (C)Y-BOCS = (Children’s) Yale-Brown Obsessive-Compulsive Scale; dlPFC = dorsolateral prefrontal cortex; L = left; Occ = occipital; PMC = primary motor cortex; R = right; SMA = supplementary motor area; SPL = superior parietal lobule.

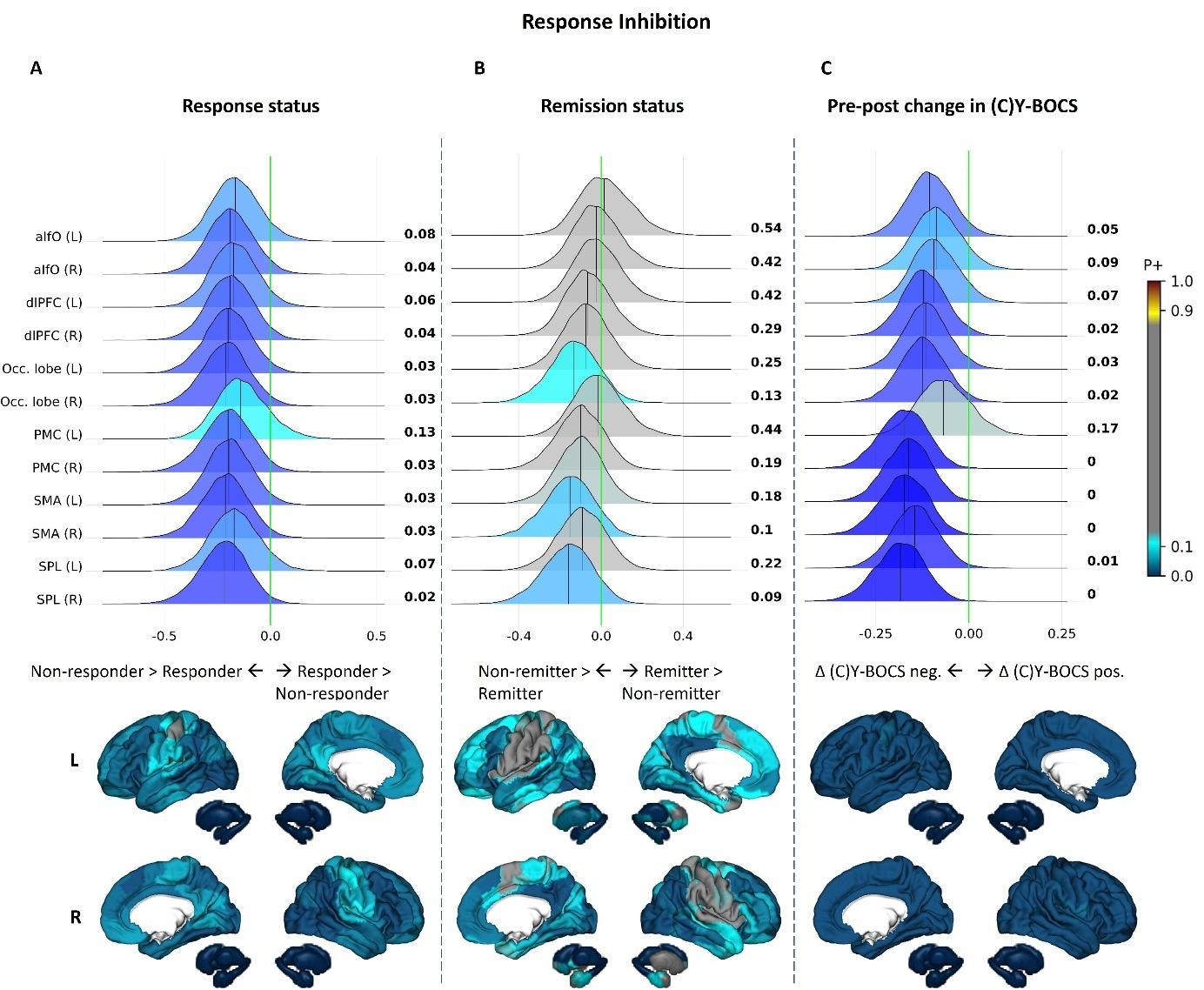

**Figure S5 – Effect of CBT outcomes on response inhibition after ComBat harmonization of sample effects** Results from Bayesian multilevel analyses of the relation between pre-treatment brain activity during response inhibition and (A) response to CBT, (B) remission after CBT, and (C) pre-to-post-CBT change in symptom severity. Sample effects were corrected for by performing ComBat harmonization on the dataset. aI/fO = anterior insula/frontal operculum; (C)Y-BOCS = (Children’s) Yale-Brown Obsessive-Compulsive Scale; dlPFC = dorsolateral prefrontal cortex; L = left; Occ = occipital; PMC = primary motor cortex; R = right; SMA = supplementary motor area; SPL = superior parietal lobule.

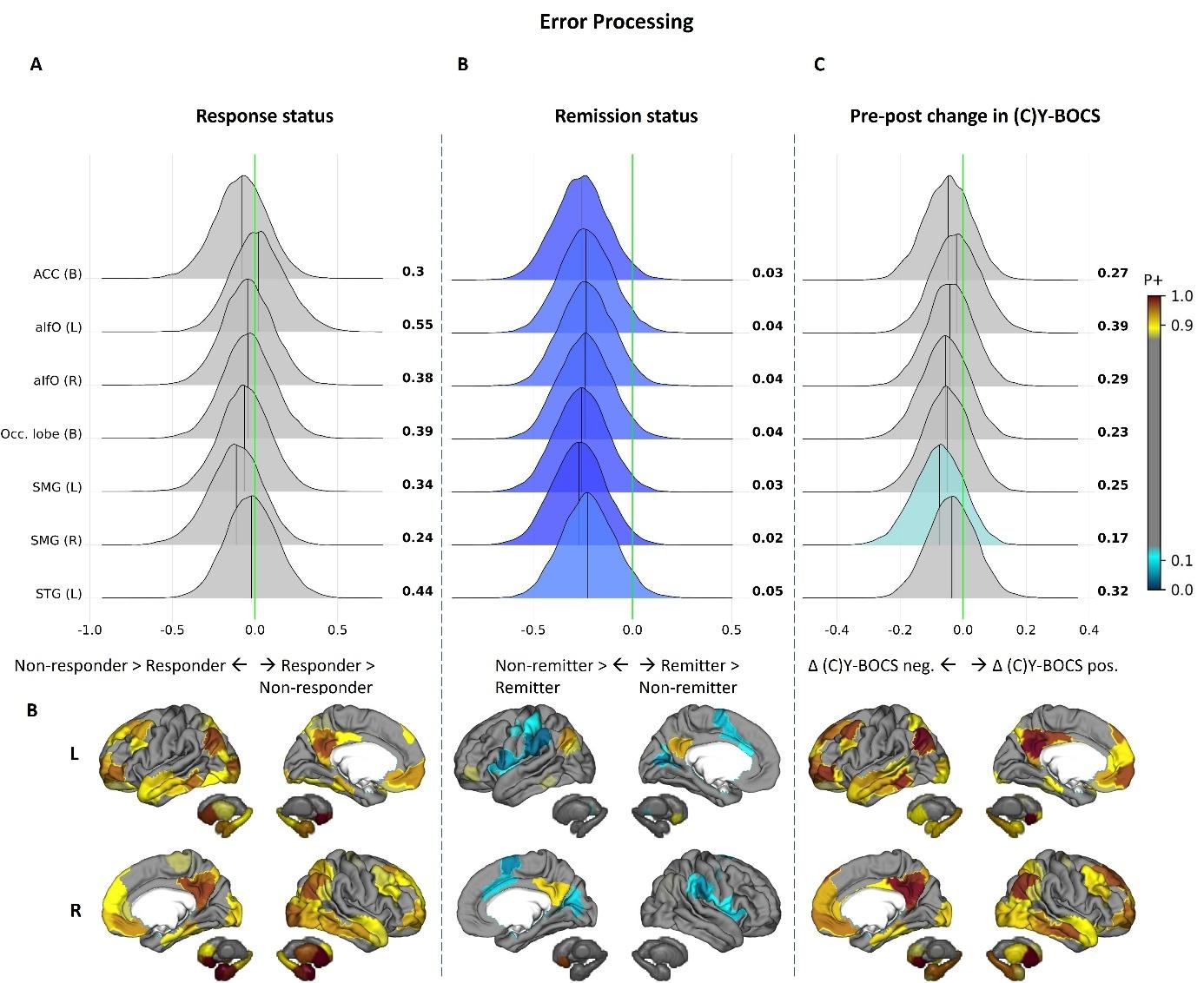

**Figure S6 – Effect of CBT outcomes on error processing** Results from Bayesian multilevel analyses of the relation between pre-treatment brain activity during error processing and (A) response to CBT, (B) remission after CBT, and (C) pre-to-post-CBT change in symptom severity. Sample effects were corrected for by including sample as a covariate of no interest. ACC = anterior cingulate cortex; aI/fO = anterior insula/frontal operculum; (C)Y-BOCS = (Children’s) Yale-Brown Obsessive-Compulsive Scale; L = left; Occ = occipital; R = right; SMG = supramarginal gyrus; STG = superior temporal gyrus.

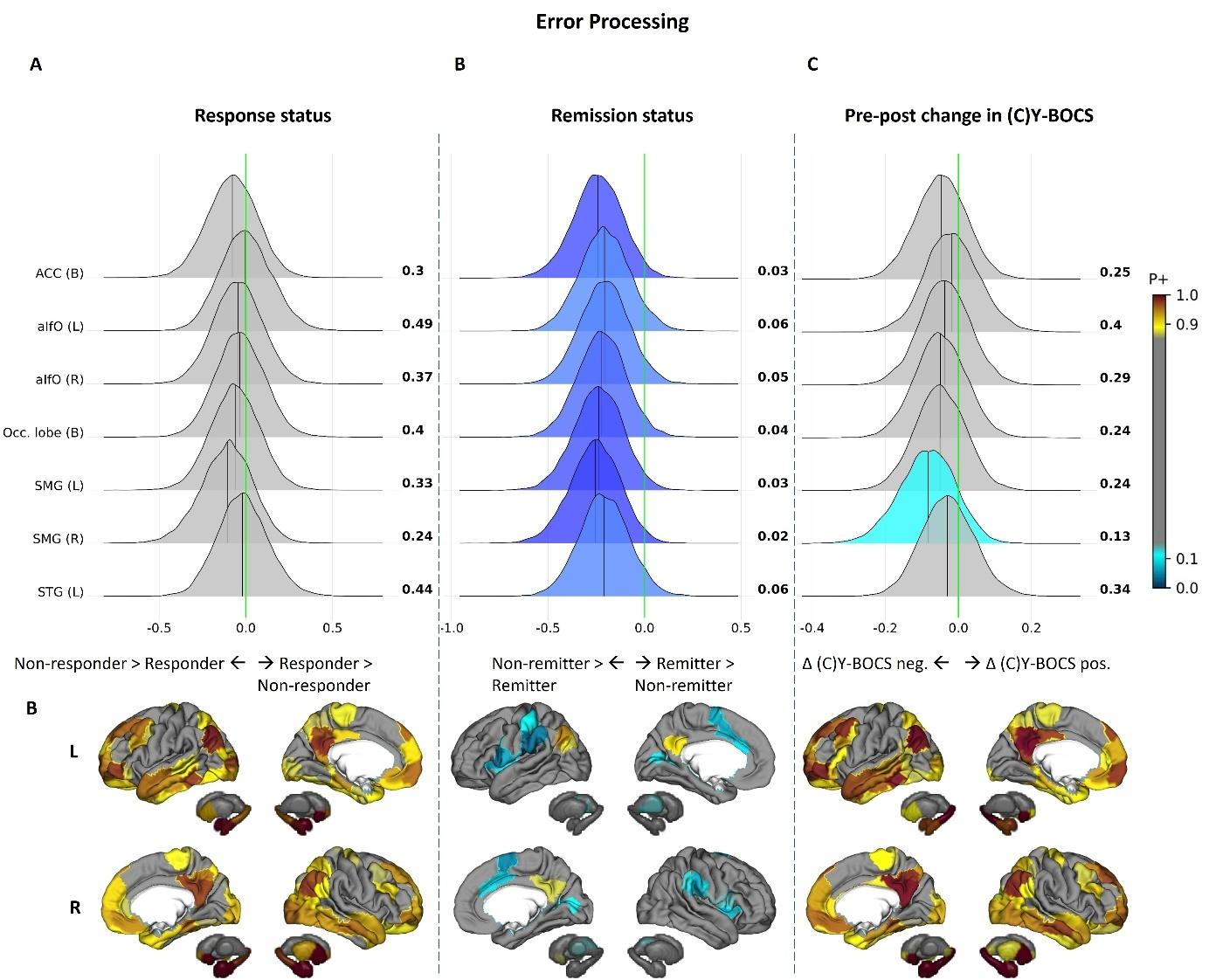

**Figure S7 – Effect of CBT outcomes on error processing after ComBat harmonization of sample effects** Results from Bayesian multilevel analyses of the relation between pre-treatment brain activity during error processing and (A) response to CBT, (B) remission after CBT, and (C) pre-to-post-CBT change in symptom severity. Sample effects were corrected for by performing ComBat harmonization on the dataset. ACC = anterior cingulate cortex; aI/fO = anterior insula/frontal operculum; (C)Y-BOCS = (Children’s) Yale-Brown Obsessive-Compulsive Scale; L = left; Occ = occipital; R = right; SMG = supramarginal gyrus; STG = superior temporal gyrus.
